# Evaluating the impact of antiviral post-exposure prophylaxis for health-care workers during ebolavirus outbreaks: a modelling study

**DOI:** 10.64898/2026.06.26.26356717

**Authors:** Jacob N. Stapley, Youngsuk Ko, Geetha Jeyapragasan, Elana M.G. Chan, Abel W. Walekhwa, Cathal Mills, Christophe Fraser, Arthur L. Reingold, Dav Ebengo, Christl A. Donnelly, Placide K. Mbala, Charles Whittaker

## Abstract

**Background:** Ebolavirus outbreaks place health-care workers (HCWs) at substantial risk, and HCW illness or death can weaken response capacity. The 2026 Bundibugyo virus disease outbreak in the DRC highlights the need for deployable countermeasures where species-specific vaccines are unavailable. With candidate antivirals under evaluation, but limited evidence on how best to use them if they become available, we assessed HCW-targeted antiviral post-exposure prophylaxis (PEP) across deployment and disruption scenarios.

**Methods:** We developed a stochastic branching-process model of ebolavirus transmission, representing health-care, community, and funeral transmission, time-varying non-pharmaceutical interventions and HCW-targeted PEP. Calibrated to previous outbreaks (a high-burden “West Africa-like” scenario and a “DRC-like” scenario with conflict-disrupted response) we estimated HCW deaths and capacity loss averted by PEP, varying antiviral efficacy, coverage, deployment readiness, dosing delay, disruption, and allocation policy.

**Findings:** At 80% efficacy and 80% coverage, PEP averted approximately 64% of HCW deaths across both archetypes. Under low readiness, where clinical evaluation and access expansion occurred during the outbreak and coverage reached only 50% over one year, reductions fell to 19% and 22% in the West Africa-like and DRC-like archetypes. In disruption scenarios, delayed dose receipt reduced HCW deaths averted from 83% to 50%; combined dosing delay and coverage loss retained only 35% of achievable impact. Dose efficiency varied by allocation: targeting recognised highest-risk exposures required 44 doses per HCW death averted, compared with 109 under broad allocation.

**Interpretation:** HCW-targeted antiviral PEP could reduce occupational mortality and preserve clinical capacity during ebolavirus epidemics where vaccines are unavailable, but realising this benefit depends on implementation readiness, rapid delivery, operational coverage and efficient allocation.

**Funding:** Gilead Sciences; UK NIHR; Oxford Martin School; Miller Institute; EDCTP; CEPI

## INTRODUCTION

The 2013–16 West African Ebola virus (EBOV; *Zaire ebolavirus*) epidemic demonstrated the high-mortality consequences of delayed detection and uncontrolled transmission of ebolaviruses^1,2^. Recent filovirus outbreaks across East Africa, including the ongoing 2026 Bundibugyo virus outbreak in the Democratic Republic of the Congo (DRC), show that this threat extends beyond EBOV^3–5^. Historically, control has relied upon non-pharmaceutical interventions (NPIs), including contact tracing, case isolation, Ebola treatment unit (ETU) access, and safe and dignified burials (SDBs). These measures reduce community, health-care-associated, and funeral-associated transmission^4,6,7^ but are difficult to implement rapidly in remote, resource-constrained, or conflict-affected settings in which many Ebola outbreaks occur^8–10^.

Medical countermeasures have substantially improved preparedness for, and response to, EBOV. The rVSV-ZEBOV vaccine has shown high efficacy when deployed through ring-vaccination strategies, protecting exposed contacts and improving outbreak-control prospects by interrupting transmission around detected cases^11,12^. Monoclonal antibody therapies have also reduced mortality among infected patients, lowering the clinical burden of Ebola virus disease in treated cases^11,13^. However, these advances have not produced equivalent protection across the full *Orthoebolavirus* genus, and EBOV remains the only ebolavirus for which licensed and approved therapeutic options exist^14,15^. For ebolaviruses other than EBOV, such as Bundibugyo virus and Sudan virus, response therefore continues to rely primarily on contact tracing, case isolation, infection prevention and control (IPC), and SDB, measures that depend on staffing, supplies, and secure access that are often limited early in ebolavirus outbreaks.

This absence of licensed countermeasures for several priority ebolaviruses has motivated calls to expand species-specific vaccines and therapeutics for known high-risk viruses and to develop broadly active filovirus antivirals^15^. Obeldesivir (ODV) is a promising example of a candidate broad-spectrum filovirus antiviral. In non-human primate studies, oral ODV used as post-exposure prophylaxis (PEP) protected against lethal challenge with Sudan virus, EBOV, and Marburg virus^16–18^. Although ODV is not licensed for human use, safety and tolerability data from COVID-19 studies^19^ and ethics approval for PEP deployment during the recent Marburg outbreak in Rwanda supported its inclusion in the EBO-PEP-BUNDI trial^20^. World Health Organization (WHO) expert groups have also prioritised MBP134, maftivimab, remdesivir, monoclonal antibody–remdesivir combination therapy, and candidate vaccines for evaluation during the ongoing 2026 Bundibugyo outbreak, although no licensed vaccine or therapeutic is currently available^15^.

If efficacious antivirals become available, the immediate operational challenge is to determine how they should be deployed during an outbreak. At present, no antiviral is approved for Ebola virus disease, and so there remains substantial uncertainty over which use cases (including clinical treatment, PEP, ring-based administration, or targeted protection of high-risk groups) would deliver greatest public-health value. This uncertainty is especially important when early supply is likely to be limited, because decisions about who receives antivirals first will determine whether they primarily reduce mortality among infected patients, interrupt transmission, protect scarce response personnel, and/or preserve health-system capacity. Health-care worker (HCW)-targeted PEP represents a compelling initial use case. HCWs are disproportionately exposed during ebolavirus outbreaks^21^, particularly early in the response before diagnosis, personal protective equipment (PPE)/IPC, and isolation systems are fully established. HCW infections remove scarce clinical staff from the response, disrupt routine health-care delivery, and can contribute to further nosocomial transmission^22,23^.

Despite this, the potential population-level impact of HCW-targeted antiviral PEP, and its dependence on efficacy, coverage, access, and allocation policy, has not been quantified across realistic ebolavirus outbreak settings. Here, we focus on settings such as the ongoing 2026 Bundibugyo outbreak, where species-specific vaccines are, as of June 2026, unavailable, delayed, or not readily deployable, and where antiviral PEP could provide an immediately actionable strategy to protect exposed HCWs. Motivated by this gap, we developed an ebolavirus-specific adaptation of an existing stochastic branching process modelling framework. We used this model to estimate the impact of HCW-targeted antiviral PEP across calibrated outbreak archetypes, varying assumed antiviral efficacy, deployment readiness, coverage, and allocation policy. We evaluated reductions in HCW deaths as the primary outcome, with HCW-days lost used as a measure of preserved response capacity.

## METHODS

### Stochastic branching-process model of ebolavirus transmission and response

We extended a previously published stochastic branching-process model to capture key features of ebolavirus epidemiology and outbreak response^24^. The model simulates individual transmission chains, classifying infections as occurring in the general population or in HCWs, and allows transmission to occur in community, health-care, or funeral settings. It represents overdispersed transmission, health-care exposure and HCW infection, and progression from infection to symptom onset, hospitalisation, recovery or death. NPIs act on their target routes: hospitalisation and care delays shape community transmission, ETU isolation and PPE reduce health-care transmission, and SDBs reduce funeral transmission. Response parameters vary over calendar time, with trajectories for hospitalisation probability, delay to hospitalisation, ETU use, PPE coverage, and SDB coverage informed by literature-derived estimates and outbreak reports. Antiviral PEP was modelled for HCWs following potentially infectious exposure, with receipt determined by scenario-specific coverage and antiviral efficacy. Among recipients, efficacy was applied before clinical progression and onward transmission. Prevented infections were therefore removed from the transmission tree and did not contribute to subsequent disease, death, HCW-days lost, or secondary infections. The model was implemented in R and is available at https://github.com/petal-code/fiber. Full details are provided in the **Appendix**.

### Model Calibration and uncertainty

We calibrated the branching-process model to two historical outbreaks using sequential approximate Bayesian computation implemented in the R package *EasyABC*. We used the 2013–2016 West Africa epidemic to define a high-burden, reasonable worst-case archetype and the 2018–2020 North Kivu and Ituri outbreak in eastern DRC, to define an archetype with prolonged transmission under conflict-related response disruption. We fitted five parameters defining each archetype: the basic reproduction number, the proportion of transmission attributable to funerals, PPE and ETU efficacy scalars, and a scalar controlling health-care setting exposure among HCWs. For each parameter set, we simulated multiple stochastic outbreaks and compared trajectories with observed targets for epidemic take-off (>100 cumulative deaths), total deaths, HCW deaths, and peak weekly deaths. The resulting posterior samples for each archetype (“West Africa-like” and “DRC-like”) were propagated into paired counterfactual simulations comparing baseline outbreaks without PEP with otherwise matched scenarios in which PEP was available.

### Simulation experiments and sensitivity analyses

For each archetype, we simulated paired outbreaks with and without HCW-targeted antiviral PEP, varying efficacy, coverage, deployment readiness, timeliness of post-exposure dosing, and allocation policy. The primary outcome was HCW deaths averted. Secondary outcomes were HCW-days lost averted and PEP doses required per HCW death averted.

### Deployment readiness scenarios

Three antiviral implementation scenarios were assessed. Scenario 1 (“high readiness, pre-positioned stockpiles”) assumed immediate 100% coverage, representing an outbreak-ready product with clinical, regulatory, delivery, manufacturing, and stockpiling systems in place. Scenario 2 (“intermediate readiness, no stockpiles”) assumed scale-up to 80% coverage over 180 days, representing a deployment-ready product with sufficient prior evidence to support outbreak use but constrained by supply. Scenario 3 (“low readiness”) assumed slower scale-up to 50% coverage over one year, representing a product with early human safety data but no outbreak-based efficacy evidence, such that access expands during concurrent clinical evaluation and regulatory decision-making.

### Effect of delayed dosing due to operational disruption

We fitted a piecewise-exponential proportional-hazards model to non-human primate survival data from prior ODV challenge studies, using time from challenge to first dose to estimate how prophylactic efficacy declines with dosing delay **(Figure S3)**. The fitted model defined a central delay–efficacy relationship, with 95% confidence intervals used as “optimistic” and “pessimistic” curves representing uncertainty in how quickly protection is lost as dosing is delayed **(Figure S4)**. We used these curves to explore how delayed receipt of antiviral PEP could reduce impact, even when coverage is high, and evaluated this mechanism in the DRC-like archetype using conflict-associated disruption as a motivating example. We defined a “maximum achievable-impact” benchmark with high PEP coverage and no delay to first dose. Against this benchmark, we modelled disruption scenarios introducing delayed dosing, reduced PEP coverage, or both, to quantify how much impact would be lost if operational conditions deteriorated. The timing and shape of disruption were informed by SDB performance data^9^ and community-death trends from WHO and UNICEF situation reports **(Table S4, Figure S5)**. For each scenario, we estimated cumulative HCW deaths and the proportion of achievable HCW deaths averted.

### Allocation policies and stockpile analyses

We evaluated how allocation policy affected PEP dose requirements for HCW protection. Under Policy A, PEP was offered broadly to all eligible HCWs following a potentially infectious exposure before accounting for whether other protective measures (e.g., PPE) would have reduced infection risk. This conservative approach is operationally simple but dose-intensive and may be limited by the safety, tolerability, pharmacokinetics and feasibility of repeated PEP courses. Under Policy B, PEP was targeted to high-risk exposures such as PPE breaches, direct body-fluid contact, or care outside an ETU. This policy represents an idealised lower bound on dose requirements but assumes that high-risk exposures can be identified and prioritised. Dose efficiency was defined as PEP doses per HCW death averted. Unless otherwise stated, reported estimates are posterior medians with 95% credible intervals.

## RESULTS

### Antiviral PEP could substantially reduce HCW deaths across diverse outbreak archetypes

We evaluated HCW-targeted antiviral PEP using a stochastic branching-process model calibrated to previous outbreaks, defining two contrasting outbreak archetypes: a high-burden “West Africa-like” reasonable worst-case setting **(Figure 1a)**, and a “DRC-like” setting in which conflict and instability disrupt response and prolong transmission **(Figure 1b)**. In the no-PEP baseline, cumulative HCW deaths reached a median of 553 (interquartile range (IQR): 208-983) in the West Africa-like archetype by week 60, compared with 61 (IQR: 19-125) in the DRC-like archetype by week 80. Assuming 80% efficacy and 80% coverage with PEP, cumulative HCW deaths fell to 200 (IQR: 68-349) and 22 (IQR: 10-44) in the West Africa-like and DRC-like archetypes, respectively, equivalent to reductions of 64% (IQR: 63-66%) and 64% (IQR: 60-68%), respectively, relative to baseline **(Figure 1c, 1d)**.

**Figure 1.**
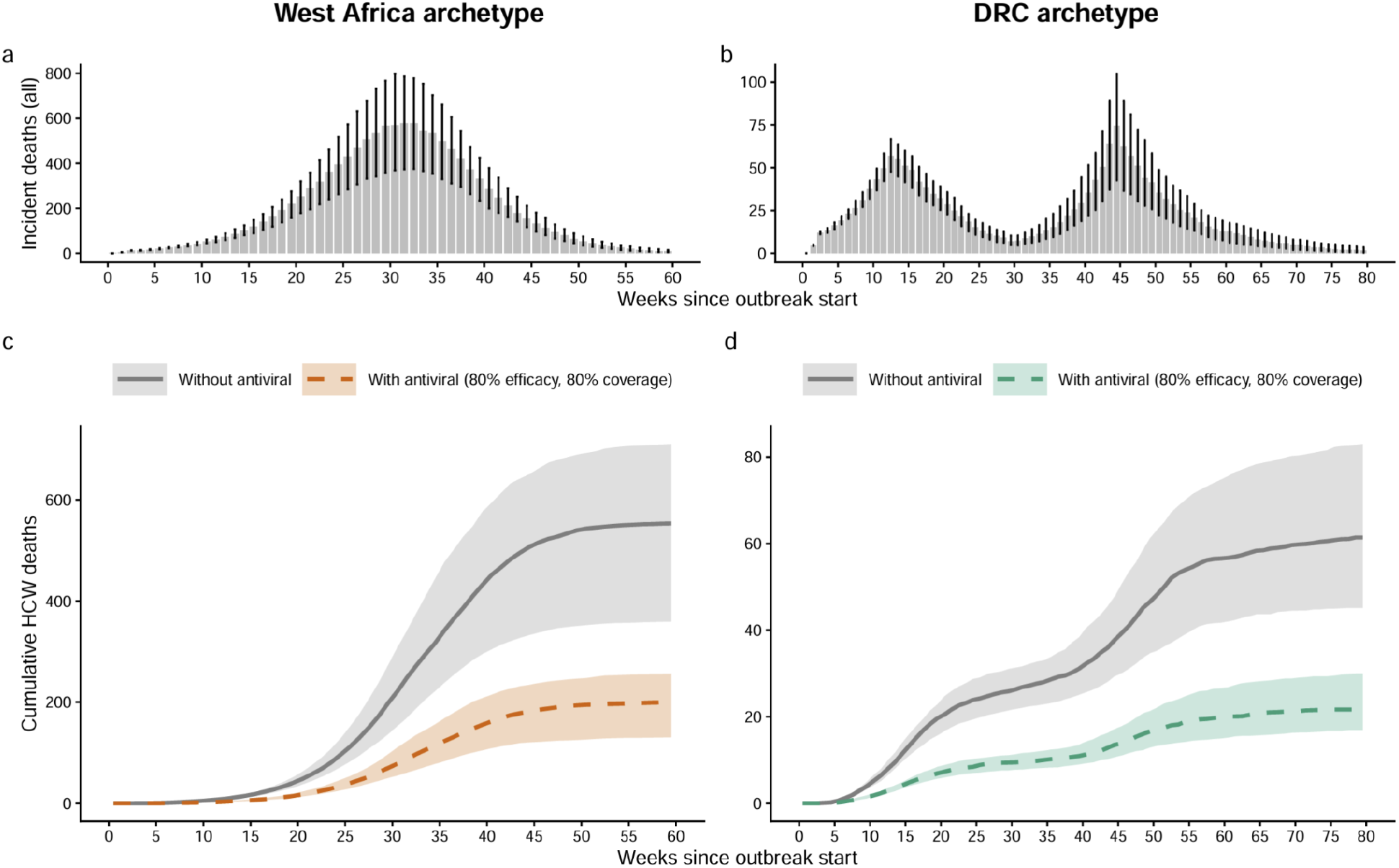
Calibration of outbreak archetypes and illustrative impact of HCW-targeted antiviral PEP. A stochastic branching-process model was calibrated to historical ebolavirus outbreak dynamics and used to define two outbreak archetypes for subsequent analyses. **a**, Weekly incident deaths for the West Africa-like archetype. **b**, Weekly incident deaths for the DRC-like archetype. Bars indicate mean simulated weekly mortality in the entire population, and vertical intervals indicate uncertainty across calibrated stochastic simulations. **c**, Cumulative HCW deaths in the West Africa-like archetype, comparing the no-PEP counterfactual with HCW-targeted antiviral PEP assuming 80% efficacy and 80% coverage. **d**, As for **c**, but for the DRC-like archetype. Grey lines/ribbons indicate simulations without PEP; coloured dashed lines/ribbons indicate simulations with PEP. Shaded regions indicate the interquartile range (25th–75th percentiles) across calibrated stochastic simulations. DRC=Democratic Republic of the Congo; HCW=health-care worker; PEP=post-exposure prophylaxis.

### Delayed antiviral availability reduced overall PEP impact on HCW burden

We examined how deployment readiness, and thus speed of antiviral availability, shaped PEP impact and compared three coverage trajectories representing distinct preparedness states (**Figure 2a–c**). Under Scenario 1, the number of HCW deaths averted increased with antiviral efficacy; at 80% efficacy, median reductions reached 80% (95% CrI: 79–81%) in the West Africa-like archetype and 80% (95% CrI: 76–84%) in the DRC-like archetype (**Figure 2f–g**). Reductions were smaller when access was scaled up during the response. Under Scenario 2, an 80%-efficacious antiviral reduced HCW deaths by 60% (95% CrI: 57–62%) and 52% (95% CrI: 41–58%) in the West Africa-like and DRC-like archetypes, preserving 75% and 66% of the Scenario 1 benefit, respectively (**Figure 2f–g**). Under Scenario 3, the corresponding reductions were 19% (95% CrI: 16–22%) and 22% (95% CrI: 7–29%) (**Figure 2f–g**). These results show that the value of antiviral PEP depends not only on antiviral efficacy, but also on deployment readiness: the evidence, stockpiling, manufacturing and delivery systems that enable courses to be distributed before HCW exposure risk has accumulated.

**Figure 2:**
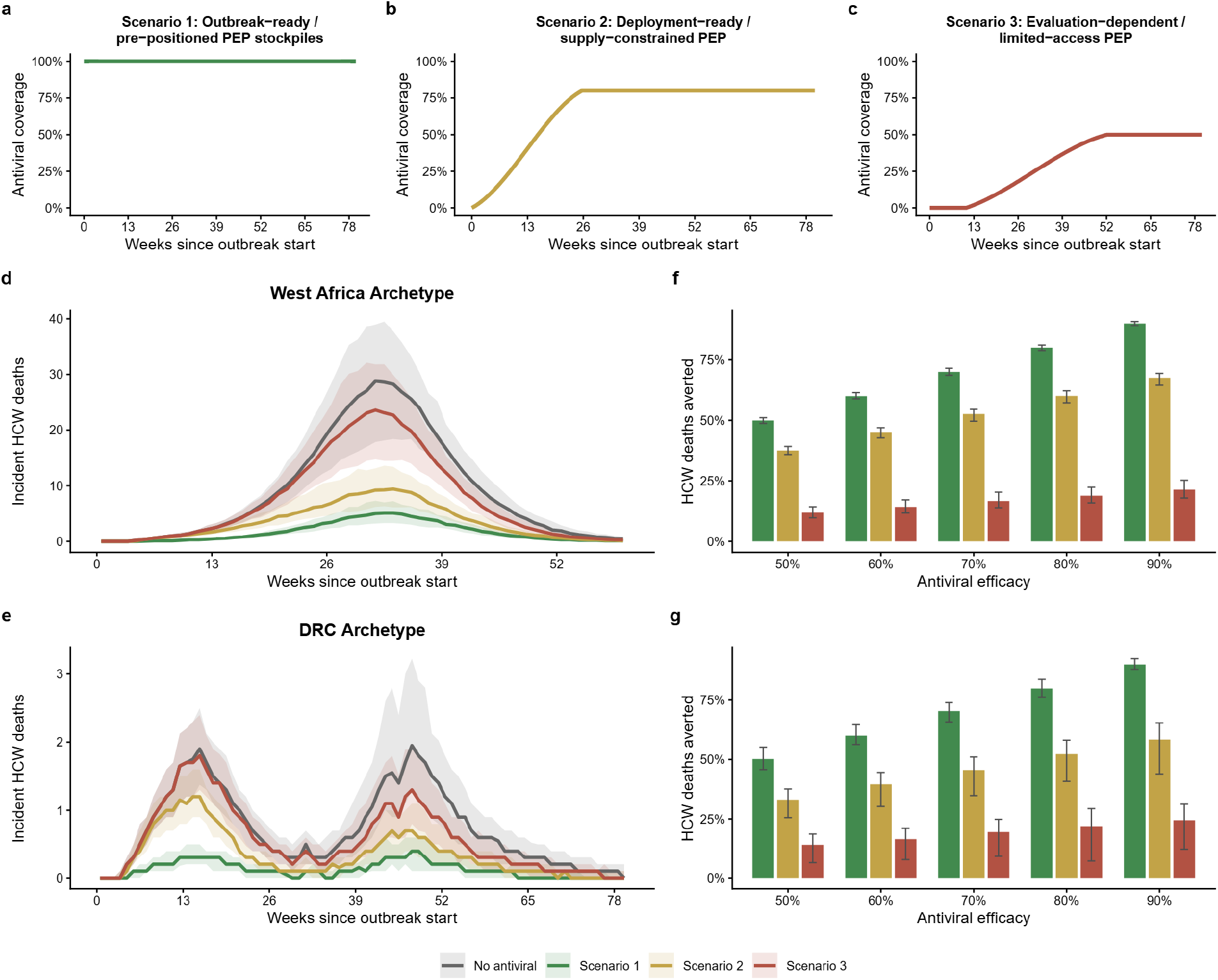
Effect of delayed PEP implementation on HCW deaths averted. Three implementation scenarios were compared to assess how timing and final coverage modify the impact of HCW-targeted antiviral PEP. **a**, Immediate full coverage, with PEP available at 100% coverage from the start of the outbreak. **b**, Gradual scale-up to high coverage, with coverage increasing from 0% to 80% over the first 180 days before plateauing. **c**, Delayed scale-up to medium coverage, with coverage remaining at 0% for the first 75 days before increasing to 50% by day 365. **d**, Weekly incident HCW deaths at 80% antiviral efficacy under each of the three coverage scenarios (no antiviral, immediate full coverage, gradual scale-up to high coverage, and delayed scale-up to medium coverage) for the West Africa-like archetype. **e**, As for **d**, for the DRC-like archetype. **f**, Percentage of HCW deaths averted under each coverage scenario and a range of assumed antiviral efficacy values for the West Africa-like outbreak archetype. **g** As for **f**, but for the DRC-like archetype. Lines and shading in **d**,**e** show medians and interquartile ranges across calibrated stochastic simulations. Lines and shading in **f**,**g** show medians and 95% credible intervals across calibrated stochastic simulations. DRC=Democratic Republic of the Congo; HCW=health-care worker; PEP=post-exposure prophylaxis.

### Operational disruption reduced the achievable impact of PEP by limiting coverage and delaying post-exposure dosing

We next examined how conflict-associated operational disruption could erode the benefit of an otherwise efficacious antiviral PEP strategy. Using a DRC-like operational-disruption scenario in which coverage among eligible HCW exposures fell and the delay from exposure to first dose increased before partial recovery of delivery capacity (**Figure S12**), we simulated four delivery scenarios and compared their impact against a no-antiviral counterfactual. In a scenario in which PEP was delivered at high coverage with no post-exposure delay (the maximum achievable impact), HCW-targeted PEP averted 83% (95% CrI: 79–87%) of HCW deaths. Introducing delayed dosing while preserving coverage reduced this to 50% (95% CrI: 40–55%) of HCW deaths averted, corresponding to only 61% (95% CrI: 49–68%) of the achievable impact retained (**Figure 3a**). When delayed dosing was combined with reduced coverage, cumulative HCW deaths increased further (**Figure 3a,b**). By decomposing impact across scenarios in which coverage, dosing delay, or both were disrupted, we partitioned the loss of achievable impact into components attributable to each mechanism (**Figure 3b**). Under optimistic, central, and pessimistic delay–efficacy assumptions, disrupted delivery realised only 36% (95% CrI: 28–48%), 35% (95% CrI: 25–47%), and 28% (95% CrI: 19–38%) of the achievable HCW deaths averted, respectively (**Figure 3c**). These results suggest that outbreak-response resilience for targeted antiviral PEP depends not only on reaching exposed HCWs, but also on initiating treatment rapidly after exposure during periods of disruption.

**Figure 3.**
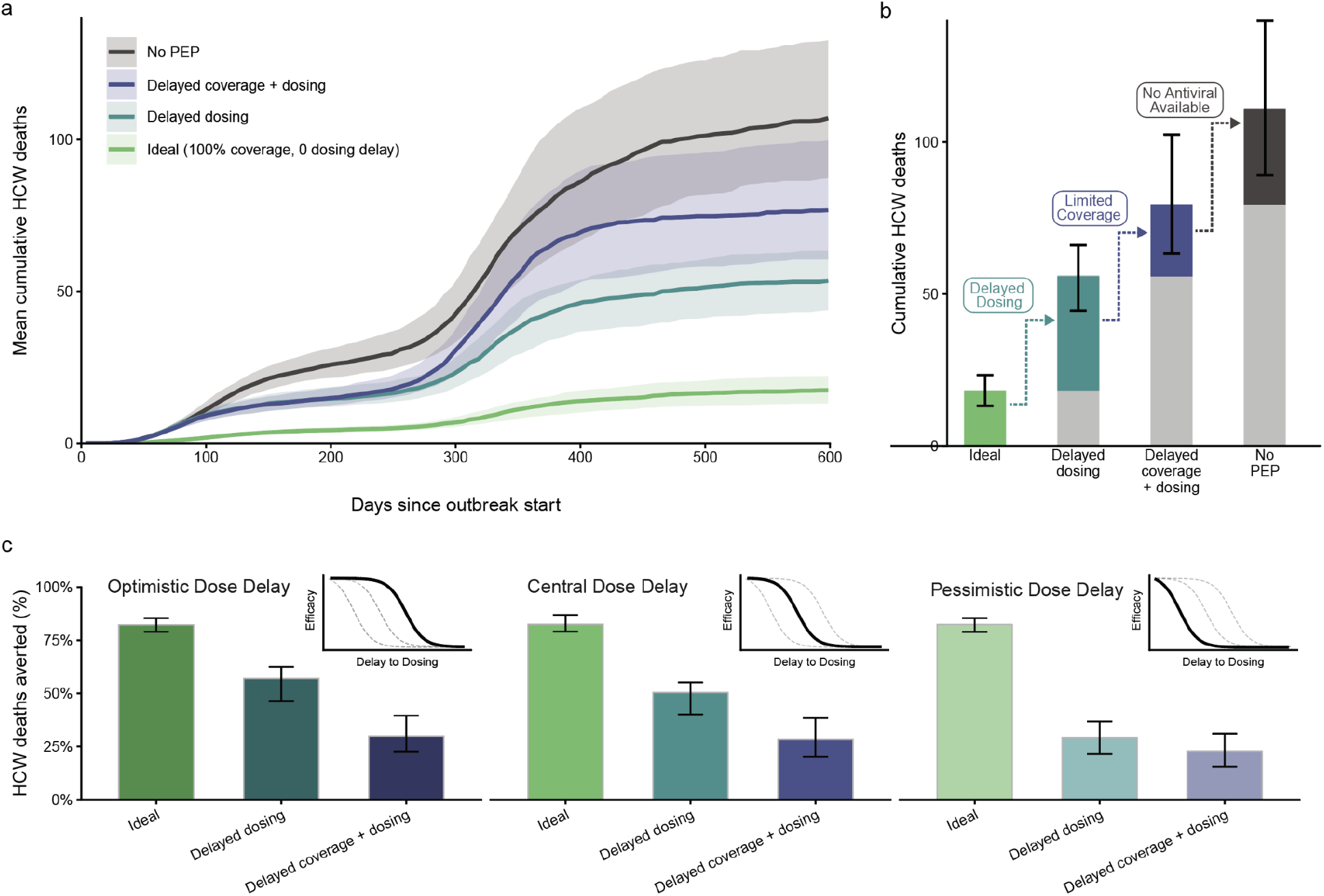
Conflict-associated disruption to coverage and dosing timeliness reduces the achievable impact of HCW-targeted antiviral PEP in the DRC-like archetype. **a**, Mean cumulative HCW deaths under four delivery scenarios (central delay–efficacy assumption): no PEP (black), ideal delivery with 100% coverage and no dosing delay (green), delayed dosing only with coverage preserved (teal), and combined coverage reduction plus dosing delay (blue). Shaded areas show the interquartile range across calibrated stochastic simulations. **b**, Cumulative HCW deaths at the end of follow-up under the same four scenarios. Bars ascend left to right as delivery conditions worsen: the green bar shows deaths under ideal delivery; the teal bar shows the additional deaths attributable to dosing delay alone; the blue bar shows the further increase attributable to reduced coverage; and the black bar shows the remaining gap to the no-antiviral counterfactual, which is shown as the combined grey and black bar on the right. Error bars indicate 95% credible intervals. **c**, Percentage of HCW deaths averted relative to the no-antiviral counterfactual for the ideal delivery, delayed dosing, and delayed coverage plus dosing scenarios, under optimistic, central, and pessimistic delay–efficacy assumptions, with inset plots showing the corresponding delay–efficacy curves. Bars show medians and error bars show 95% credible intervals across calibrated stochastic simulations. DRC=Democratic Republic of the Congo; HCW=health-care worker; PEP=post-exposure prophylaxis.

### Allocation strategy and delivery timing strongly shaped dose requirements for HCW protection

Lastly, we evaluated how stockpile size, allocation policy, and delivery timing affected the number of antiviral doses required to protect HCWs. We compared broad allocation to all eligible HCW exposures (Policy A) with targeted allocation to high-risk exposures, such as PPE breaches or direct body-fluid contact (Policy B). Expressing supply relative to targeted demand showed that Policy B reached near-maximal impact once available supply was sufficient to cover high-risk exposures, whereas Policy A required approximately 2.5-fold more doses to achieve a similar proportion of HCW deaths averted **(Figure 4a)**. Across both archetypes, HCW deaths averted rose with stockpile size before plateauing once available doses covered eligible exposures **(Figure 4b,c)**. Delivery timing also strongly affected impact. At stockpiles of 30,000 doses in the West Africa-like archetype and 2,500 doses in the DRC-like archetype, prompt delivery averted 466 and 29 HCW deaths, respectively. Delaying the first dose to 5 days post-exposure reduced these benefits by 164 and 10 deaths, respectively. For comparable reductions in HCW deaths, dose-efficiency analyses showed that doses required per HCW death averted were consistently higher when delivery was delayed and when PEP was allocated broadly **(Figure 4d)**. At 80% antiviral efficacy with same-day dosing, targeted allocation required 44 doses per HCW death averted, compared with 109 doses under broad allocation. With delayed delivery, dose requirements increased to 68 and 168 doses per HCW death averted under targeted and broad allocation, respectively. Together, these results indicate that antiviral PEP stockpile planning should consider not only the number of doses available, but also how rapidly courses can be delivered after exposure and how efficiently they can be prioritised when supply is limited.

**Figure 4.**
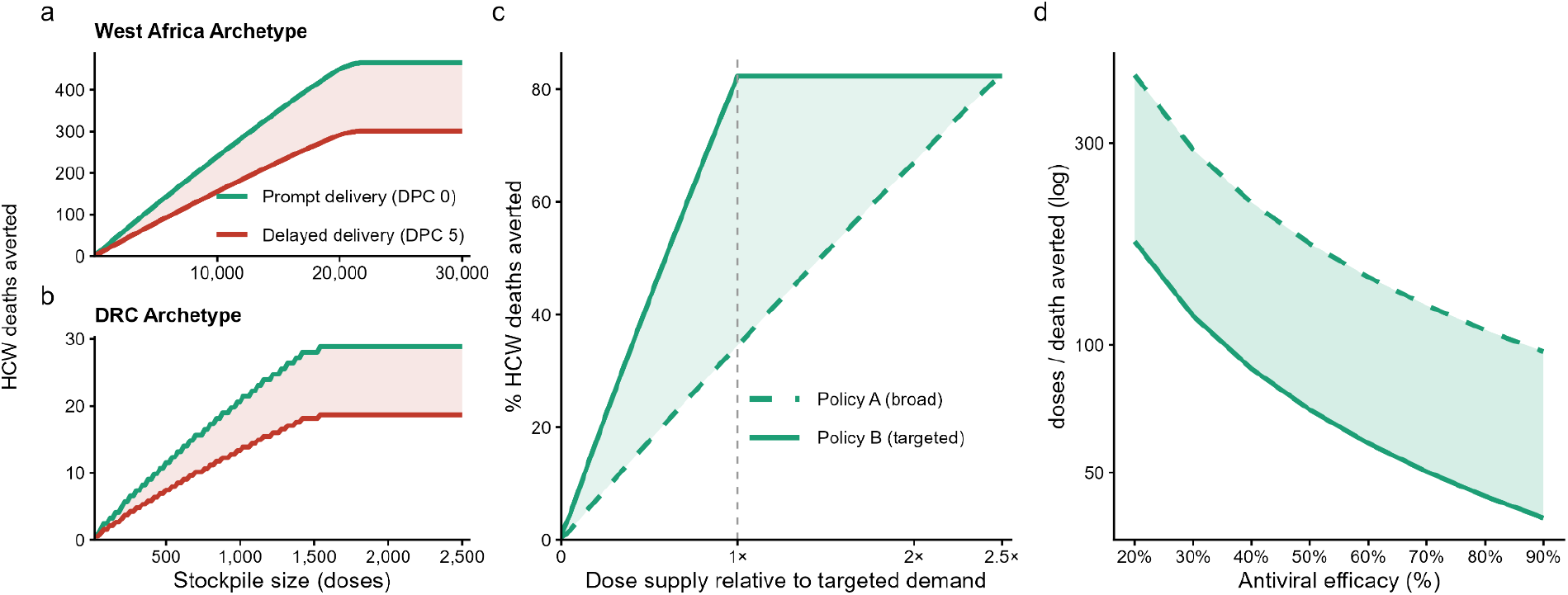
Dose requirements for HCW protection depended on allocation strategy, stockpile size, and delivery timing. **a**, HCW deaths averted as a function of finite stockpile size for the West Africa-like archetype under targeted allocation, comparing prompt delivery with same-day dosing after exposure (DPC 0; green) and delayed delivery with first dose 5 days after exposure (DPC 5; red). **b**, As for a but for the DRC-like archetype. **c**, Percentage of HCW deaths averted as a function of available supply, expressed relative to targeted demand, for the West Africa-like archetype. Solid line shows targeted allocation to high-risk exposures (Policy B), while dashed line shows broad allocation to all eligible HCW exposures (Policy A). The dashed vertical line indicates supply equal to targeted demand (1×). **d**, Doses required per HCW death averted as a function of intrinsic antiviral efficacy, shown on the log scale. Solid lines indicate targeted allocation (Policy B), dashed lines indicate broad allocation (Policy A). DPC=days post-exposure to first dose; DRC=Democratic Republic of the Congo; HCW=health-care worker; PEP=post-exposure prophylaxis.

## DISCUSSION

Our results suggest that antiviral PEP could substantially reduce HCW deaths during ebolavirus outbreaks in settings where species-specific vaccines are unavailable, delayed, or not readily deployable, such as the ongoing 2026 Bundibugyo outbreak. However, impact depended strongly on whether PEP could be delivered early, consistently, and to the right exposures. Delayed availability reduced impact even when eventual coverage was high; operational disruption further eroded achievable benefit by delaying first dose after exposure and reducing coverage among eligible HCWs. Dose requirements were also shaped by delivery timing and allocation strategy: broad allocation and delayed delivery required substantially more doses per HCW death averted than prompt, targeted allocation, reducing the number of HCW deaths that could be averted from any fixed stockpile of doses. Broad allocation reduces the risk of missing exposed HCWs but may be constrained by repeated-course safety or pharmacology, whereas targeted allocation improves dose efficiency but depends on accurate exposure recognition. These results show that PEP value depends not just on antiviral efficacy, but also deployment readiness, dosing timeliness, coverage, and allocation efficiency.

The potential impact of HCW-targeted PEP is important because HCWs are both a high-risk group and a core component of the outbreak response. HCW infection and mortality have been recurrent features of ebolavirus outbreaks, particularly early in response operations when diagnosis is delayed, PPE and IPC practices are still being established, and ETU access is limited^21–23^. Reducing HCW infection and death is therefore important both for occupational protection and for preserving the workforce needed to provide care, staff ETUs, and support the response. Preserving HCW capacity is especially relevant where trained clinical staff are scarce, because HCW illness and death can weaken outbreak response and routine health services^22^.

Achieving this impact will require more than an efficacious antiviral; enabling infrastructure for rapid evaluation, deployment, equitable access, and timely post-exposure delivery will also be needed. PEP can only prevent infections that have not yet occurred, so both the timing of product availability and the delay from exposure to first dose shape impact. Across the deployment-readiness scenarios we explored, the largest reductions in HCW deaths occurred when PEP was available from the start of the outbreak, consistent with a product for which clinical evaluation, regulatory and ethical pathways, delivery protocols, manufacturing capacity, and stockpiles are in place before outbreak detection. Scenarios requiring manufacturing scale-up or concurrent outbreak-linked clinical evaluation achieved substantially smaller reductions, even when the assumed antiviral efficacy was unchanged. Our analyses exploring operational disruption further underscore this point: even when drug supply exists, conflict-associated disruption can reduce realised impact if exposed HCWs receive PEP late or if coverage falls during periods of instability. During the ongoing 2026 Bundibugyo outbreak in DRC and Uganda, WHO expert groups have prioritised several products for evaluation on compressed timelines^25^, reflecting prior investment in trial protocols, regulatory and ethical pathways, safety monitoring, manufacturing, procurement, stockpiling, and delivery systems^26^. Our results underscore the value of these investments, which determine whether a promising antiviral can reach exposed HCWs while infection can still be prevented.

Although this study explored the potential impact of antiviral PEP for HCWs, candidate filovirus antivirals could support several other outbreak-response strategies, including case treatment, PEP for household members or other high-risk contacts, ring-based administration, or prophylaxis for burial teams and informal caregivers. Crucially, these strategies differ in both their operational requirements (including dose requirements, speed of delivery after exposure, and ability to prioritise high-risk contacts) and public-health goals (reducing mortality, preventing onward transmission, preserving response capacity, etc.). Current and planned trials will provide essential evidence on safety, efficacy, and implementation, including for household PEP^20^, which is a compelling strategy given high secondary attack rates^27^ among close contacts and the contribution of household transmission to ebolavirus spread^28^. Future work should compare use cases directly, combining trial data with modelling where empirical evaluation is constrained, to identify how limited antiviral supplies can best reduce disease burden, preserve response capacity, and support containment.

Our study has several limitations. First, no antiviral is currently approved for Ebola virus disease, and the efficacy, adherence, timing, and clinical effects of candidate products in human filovirus exposures remain uncertain. Indeed, evidence for candidate antivirals against ebolaviruses is still being generated^15,20,25^. Whilst we mitigate this uncertainty in part by exploring potential impact across a range of efficacy and coverage values, realised impact will depend on both the true prophylactic effectiveness and factors that determine real-world delivery, including drug availability, recognition of eligible exposures, the interval between exposure and first dose delivery, and adherence. Relatedly, the NHP-derived delay–efficacy curves require cautious interpretation as lethal NHP challenge models have shorter incubation periods than natural human exposures. The same numerical delay may therefore correspond to a later stage of infection in NHPs than in exposed HCWs. Our optimistic delay–efficacy scenario partly captures the possibility that efficacy declines more slowly in humans because of this longer incubation period. Similarly, our operational-disruption scenarios are stylised and intended to represent how delays and coverage loss could erode impact, rather than to reconstruct a specific future conflict trajectory.

A further limitation is that we did not explicitly model ring vaccination. This reflects the primary policy question addressed here, which is the potential role of antiviral PEP in outbreaks where species-specific vaccines are unavailable, delayed, or not deployable, as is currently the case for Bundibugyo virus disease. However, it limits the interpretation of the DRC-like calibration, because rVSV-ZEBOV vaccination was deployed during the 2018–2020 North Kivu and Ituri outbreak^12^. Although sensitivity analyses **(Figure S11, S12)**, in which baseline transmissibility in the DRC-like archetype was increased to approximate vaccine-unavailable settings, showed that PEP impact was robust to higher underlying transmission, these analyses do not replace an explicit mechanistic representation of ring vaccination. The DRC-like archetype should therefore be interpreted as reproducing realised conflict-disrupted outbreak dynamics, rather than decomposing the separate effects of vaccination, NPIs, and care. More broadly, ebolavirus epidemics are highly stochastic and context-specific, and many introductions do not generate large epidemics when detection and response occur promptly^29,30^. These analyses should therefore be interpreted as scenario-based evaluations, not outbreak-specific predictions. Nevertheless, by spanning severe but plausible reasonable-worst-case outbreak conditions, they support the idea that HCW-targeted PEP could be impactful across a range of ebolavirus epidemic contexts.

Despite these limitations, our results highlight the potential impact of HCW-targeted antiviral PEP in reducing occupational mortality and preserving clinical response capacity during ebolavirus outbreaks. The greatest benefits were achieved when PEP was available rapidly, delivered at high coverage, initiated soon after exposure, and allocated efficiently. The value of PEP therefore depends not only on biological effectiveness, but on trial readiness, regulatory pathways, stockpiling, manufacturing capacity, exposure-recognition systems, and delivery pathways. Planning, development and support for these systems in advance of outbreaks will be central to realising the potential benefit of candidate antivirals for outbreak response.

## Contributors

**JNS:** Conceptualisation; data curation; formal analysis; investigation; methodology; software; validation; visualisation; writing - original draft; writing - review & editing. **YK:** Investigation; data curation; formal analysis; methodology; software; validation; visualisation; writing - review & editing. **GJ:** Formal analysis; methodology; software; validation; visualisation; writing - review & editing. **EMGC**: visualisation; writing - review & editing. **AWW**: validation; writing - review & editing. **CM**: writing - review & editing. **CAD**: methodology; validation; writing - review & editing. **ALR**: funding acquisition; writing - review & editing. **CF:** writing - review & editing. **DE**: Validation; writing - review & editing. **PM**: Supervision; writing - review & editing. **CW**: Conceptualisation; data curation; formal analysis; funding acquisition; investigation; methodology; project administration; resources; software, supervision; validation; visualisation; writing – original draft; and writing – review & editing. All authors had access to all the data in the study and all accepted responsibility for the decision to submit for publication. All authors contributed to the review and editing of the manuscript, validation and visualisation of the data, and saw and approved the final text.

## Data sharing

No identifiable human participant data were analysed; ethical approval was not required. The data used in this modelling study were derived from published reports, publicly available outbreak summaries, and reconstructed non-human primate survival data described in the Appendix. Code and processed input files required to reproduce the analyses are available at: https://github.com/petal-code/antiviral_pep_hcw_paper.

## Declaration of Generative AI and AI-assisted technologies in the writing process

During the preparation of this work the author(s) used Claude AI in order to improve clarity and conciseness of the writing. After using this tool/service, the author(s) reviewed and edited the content as needed and take(s) full responsibility for the content of the publication.

## Acknowledgments

We thank all the health professionals who are actively engaged in controlling the ongoing 2026 Bundibugyo outbreak. This study was supported by a gift from Gilead Sciences to the UC Berkeley School of Public Health (**JNS, YK, GJ, EMGC, ALR, CW**). This study was further supported by grant funding (awarded to **CAD**) from the UK National Institute for Health and Care Research (Health Protection Research Unit in Emerging and Zoonotic Infections) (grant no: HPRU200907) and the Oxford Martin School (Programme in Digital Pandemic Preparedness). **CAD** serves for the World Health Organization R&D Blueprint in an unpaid position developing and evaluating possible vaccine and therapy effectiveness study designs. **CAD** thanks the Miller Institute for Basic Research in Science, University of California, Berkeley for support in the form of a Visiting Miller Professorship. **DE** wishes to thank the European Union – Horizon Europe / EDCTP3 Joint Undertaking for supporting this work through the EBOLA PREP TBOX project (Grant Agreement No. 101145709). This work was supported by grant funding (awarded to **CF**) from CEPI for the PRESTO project.

## References

1 WHO Ebola Response Team, Aylward B, Barboza P, et al. Ebola virus disease in West Africa--the first 9 months of the epidemic and forward projections. N Engl J Med 2014; 371: 1481–95.

2 Bell BP, Damon IK, Jernigan DB, et al. Overview, control strategies, and lessons learned in the CDC response to the 2014-2016 Ebola epidemic. MMWR Suppl 2016; 65: 4–11.

3 Marburg virus disease-Rwanda. https://www.who.int/emergencies/disease-outbreak-news/item/2024-DON548? (accessed June 12, 2026).

4 Sudan virus disease – Uganda. https://www.who.int/emergencies/disease-outbreak-news/item/2025-DON558? (accessed June 12, 2026).

5 Epidemic of Ebola Disease caused by Bundibugyo virus in the Democratic Republic of the Congo and Uganda determined a public health emergency of international concern. https://www.who.int/news/item/17-05-2026-epidemic-of-ebola-disease-in-the-democratic-republic-of-the-congo-and-uganda-determined-a-public-health-emergency-of-international-concern? (accessed June 12, 2026).

6 Legrand J, Grais RF, Boelle PY, Valleron AJ, Flahault A. Understanding the dynamics of Ebola epidemics. Epidemiol Infect 2007; 135: 610–21.

7 Tiffany A, Dalziel BD, Kagume Njenge H, et al. Estimating the number of secondary Ebola cases resulting from an unsafe burial and risk factors for transmission during the West Africa Ebola epidemic. PLoS Negl Trop Dis 2017; 11: e0005491.

8 Vinck P, Pham PN, Bindu KK, Bedford J, Nilles EJ. Institutional trust and misinformation in the response to the 2018-19 Ebola outbreak in North Kivu, DR Congo: a population-based survey. Lancet Infect Dis 2019; 19: 529–36.

9 Warsame A, Eamer G, Kai A, et al. Performance of a safe and dignified burial intervention during an Ebola epidemic in the eastern Democratic Republic of the Congo, 2018-2019. BMC Med 2023; 21: 484.

10 Checchi F, Eamer G, Katshitshi J, Robles Dios L, Kai A, Warsame A. Effect of a safe and dignified burial intervention on Ebola virus transmission in the eastern Democratic Republic of the Congo, 2018-19: a propensity score analysis. Lancet Glob Health 2025; 13: e1617–26.

11 Henao-Restrepo AM, Camacho A, Longini IM, et al. Efficacy and effectiveness of an rVSV-vectored vaccine in preventing Ebola virus disease: final results from the Guinea ring vaccination, open-label, cluster-randomised trial (Ebola Ça Suffit!). Lancet 2017; 389: 505–18.

12 Muyembe J-J, Pan H, Peto R, et al. Ebola outbreak response in the DRC with rVSV-ZEBOV-GP ring vaccination. N Engl J Med 2024; 391: 2327–36.

13 Mulangu S, Dodd LE, Davey RT Jr, et al. A randomized, controlled trial of Ebola virus disease therapeutics. N Engl J Med 2019; 381: 2293–303.

14 Fischer WA 2nd, Vetter P, Bausch DG, et al. Ebola virus disease: an update on post-exposure prophylaxis. Lancet Infect Dis 2018; 18: e183–92.

15 Filovirus research and development roadmap. https://www.who.int/publications/m/item/filovirus-research-and-development-roadmap (accessed June 12, 2026).

16 Cross RW, Woolsey C, Chu VC, et al. Oral administration of obeldesivir protects nonhuman primates against Sudan ebolavirus. Science 2024; 383: eadk6176.

17 Woolsey C, Cross RW, Chu VC, et al. The oral drug obeldesivir protects nonhuman primates against lethal Ebola virus infection. Sci Adv 2025; 11: eadw0659.

18 Cross RW, Woolsey C, Prasad AN, et al. Oral obeldesivir provides postexposure protection against Marburg virus in nonhuman primates. Nat Med 2025; 31: 1303–11.

19 Ogbuagu O, Goldman JD, Gottlieb RL, et al. Efficacy and safety of obeldesivir in low-risk, non-hospitalised patients with COVID-19 (OAKTREE): a phase 3, randomised, double-blind, placebo-controlled study. Lancet Infect Dis 2025; 25: 1282–92.

20 Ebola Zaïre post-exposure prophylaxis, preparedness and efficacy evaluation during outbreak in Central and West-Africa. 2026. https://cdn.who.int/media/docs/default-source/consultation-rdb/6_ebopep.pdf?sfvrsn=6326ba1d_1.

21 Kilmarx PH, Clarke KR, Dietz PM, et al. Ebola virus disease in health care workers--Sierra Leone, 2014. MMWR Morb Mortal Wkly Rep 2014; 63: 1168–71.

22 Evans DK, Goldstein M, Popova A. Health-care worker mortality and the legacy of the Ebola epidemic. Lancet Glob Health 2015; 3: e439–40.

23 Selvaraj SA, Lee KE, Harrell M, Ivanov I, Allegranzi B. Infection rates and risk factors for infection among health workers during Ebola and Marburg virus outbreaks: A systematic review. J Infect Dis 2018; 218: S679–89.

24 Whittaker C, Barnsley G, Mesa DO, et al. Quantifying the impact of a broadly protective sarbecovirus vaccine in a future SARS-X pandemic. Nat Commun 2025; 16: 8495.

25 WHO Technical Advisory Group on therapeutics prioritization for Bundibugyo virus disease: meeting report, 20 and 26 May 2026. 2026; published online May 28. https://www.who.int/publications/i/item/B09767 (accessed June 13, 2026).

26 Saville M, Cramer JP, Downham M, et al. Delivering pandemic vaccines in 100 days -what will it take? N Engl J Med 2022; 387: e3.

27 Glynn JR, Bower H, Johnson S, et al. Variability in intrahousehold transmission of Ebola virus, and estimation of the household secondary attack rate. J Infect Dis 2018; 217: 232–7.

28 Faye O, Boëlle P-Y, Heleze E, et al. Chains of transmission and control of Ebola virus disease in Conakry, Guinea, in 2014: an observational study. Lancet Infect Dis 2015; 15: 320–6.

29 Glennon EE, Jephcott FL, Restif O, Wood JLN. Estimating undetected Ebola spillovers. PLoS Negl Trop Dis 2019; 13: e0007428.

30 Polonsky J, Mboussou F, Haskew C, De Waroux OLP, Impouma B. Lessons learnt from Ebola virus disease surveillance in Équateur Province, May–July 2018. Wkly Epidemiol Rec 2019; 94: 23–7.

