## Appendix for "Evaluating the impact of antiviral post-exposure prophylaxis for health-care workers during ebolavirus outbreaks: a modelling study"

### a1. Supplementary Methods: Branching Process Framework

#### 1.1 Overview of Stochastic Branching Process Modelling Framework

We extended a stochastic branching-process modelling framework previously developed to simulate outbreak containment of emerging infectious diseases and adapted it to represent in detail the specific epidemiological features, transmission dynamics and control measures associated with ebolavirus outbreaks[1–3]. We implement this extended framework as the open-source R package *fiber*, and use it to quantify how interventions, and in particular post-exposure prophylaxis with antivirals, alter outbreak size and the burden of infection borne by health-care workers (HCWs). The framework incorporates several features that are central to the epidemiology of ebolavirus. We distinguish two classes of infected individuals, members of the general population and HCWs, allowing the model to track the disproportionate infection and burden of ebolavirus typically associated with HCWs and the extensive nosocomial transmission that can occur during outbreaks[4,5]. Within the modelling framework, transmission can occur in any of three distinct settings. These are i) the community; ii) health-care settings, and iii) at funerals, reflecting the central role that hospital-based exposure and unsafe burial practices have played in past ebolavirus outbreaks[6]. Each infection is followed through an explicit clinical pathway, from infection through symptom onset, possible hospital admission (with the probability of hospitalisation and the delay between symptom onset and hospitalisation varying across the course of the outbreak), and death or recovery, with the case fatality risk allowed to differ between cases managed in the community and those admitted to care. The framework also represents a set of non-pharmaceutical interventions (NPIs) whose coverage can change as the response matures over the course of an outbreak: these include the isolation of cases within Ebola Treatment Units (ETU), the availability of personal protective equipment (PPE) and infection prevention and control (IPC) for HCWs, and safe and dignified burials (SDBs) for the deceased. Alongside these NPIs, we model post-exposure prophylaxis (PEP) with orally available antivirals, as a distinct pharmaceutical intervention[7,8]. A model schematic is presented in **Figure S1**. Below we describe the framework in detail in mathematical and technical terms, before detailing the implementation of post-exposure prophylaxis specifically. Code implementing the model is available at https://github.com/petal-code/fiber.

#### 1.2 Modelling ebolavirus Outbreak Dynamics

**Overview:** We model ebolavirus transmission as an individual-based, asynchronous stochastic branching process that propagates a transmission tree forward through time, one infectious case at a time. At each step, the earliest infection whose onward transmission has not yet been simulated is selected to act as an infector (a "parent"), and the secondary infections it generates (its "offspring") are sampled and added to the tree, each with an explicit infection time. Because cases are processed in order of their infection time rather than in discrete, synchronous generations, the absolute timing of every transmission event, hospital admission, and death is tracked throughout, ensuring that time-varying parameters and interventions act at the correct point in the outbreak. Each infection is assigned to one of two host classes, the general population or HCWs, and every transmission event occurs in one of three settings: i) the community; ii) health-care settings; and iii) at funerals. The number and timing of an infector's secondary infections are governed by setting- and class-specific transmissibility and generation-interval distributions, while NPIs act by reducing transmission within health-care and funeral settings. Both the coverage of these interventions and the operational parameters governing access to care, specifically the probability that a case is hospitalised and the delay from symptom onset to admission, are permitted to vary over calendar time, representing the strengthening of the outbreak response as it matures. The simulation proceeds until no further infections remain to be processed and the outbreak has ended and returns the complete transmission tree from which we derive outbreak size and the burden of infection (and mortality) among HCWs.

**Step 1: Selecting the next infection and generating its secondary infections:** From $N$ initialised seeding infections, the outbreak is propagated forward by repeatedly selecting the next case to simulate transmission for, and generating the secondary infections it produces. Because transmission is simulated asynchronously, in order of infection time, at each iteration we select the earliest infection whose onward transmission has not yet been generated. This case acts as the infector, or "parent," for the current iteration, and its infection time anchors the timing of all the events that follow. For a parent of class *j* (general population or HCW), the potential number of secondary infections it generates through community and health-care setting transmission, before any intervention is applied, is drawn from a negative binomial distribution:

$$N \sim \mathrm{NegBin}(\mathrm{mean} = R_j(t_{\mathrm{inf}}),\ \mathrm{size} = k_j)$$

Here $R_j(t_{inf})$ is the class-specific transmissibility, equal to the mean number of secondary infections produced by an infector of class j, evaluated at the parent's infection time $t_{inf}$ so that it may vary over the course of the outbreak. The size parameter $k_j$ governs the degree of overdispersion in the number of secondary infections, with smaller values corresponding to greater individual-level variation in transmission, reproducing the overdispersion and superspreading that characterise ebolavirus, in which a minority of cases are responsible for a disproportionate share of onward infection[9]. Each of the *N* potential secondary infections is then assigned a time of infection relative to the parent, drawn from a Gamma-distributed generation interval truncated to the parent's infectious period:

$$\tau \sim \mathrm{Gamma}(\alpha_j, \beta_j), \qquad \tau \in [0,\, T_{\mathrm{out}}],$$

with mean generation interval $\alpha_j/\beta_j$. Truncation at $T_{out}$, the interval from the parent's infection to their own outcome (either recovery or death), reflects that transmission through these settings ceases once a case is no longer infectious; transmission arising after death, at funerals, is treated separately (see **Step 4**). The count drawn here is the potential, pre-intervention number of secondary infections; their settings, host classes, and the reductions achieved by interventions are resolved in the steps that follow to give the realised secondary infections.

**Step 2: Assigning the setting and host class of each secondary infection:** Once a parent's potential secondary infections have been generated and timed, each is assigned the setting in which transmission occurs and the host class of the newly infected individual. The setting depends on the parent's class and on the timing of each secondary infection relative to the parent's own hospital admission. Let $\tau_k$ denote the infection time of secondary infection $k$ (relative to the parent's infection, from Step 1), and let $A$ denote the interval from the parent's infection to their admission to hospital, with $A = \infty$ for parents who are never admitted, including those whose outcome precedes admission. A general-population parent transmits in the community until admitted and in the hospital thereafter:

$$s_k = \begin{cases} \text{community}, & \tau_k < A, \\ \text{hospital}, & \tau_k \geq A. \end{cases}$$

We assume that an infected (but not yet necessarily symptomatic to the point of being unable to work and requiring hospitalisation) HCW, by contrast, continues to work until their own admission, and so has potential to generate part of their pre-admission transmission within the hospital (this fraction is typically small in practice). Each pre-admission infection ($\tau_k < A$) is therefore assigned to the hospital with probability $\rho$, the probability that a still-working infected HCW's transmission occurs in a health-care setting, and to the community otherwise; all post-admission infections ($\tau_k \geq A$) occur in the hospital:

$$\tau_k < A: \quad s_k = \text{hospital with probability } \rho,\ \text{community otherwise}; \qquad \tau_k \geq A: \quad s_k = \text{hospital}.$$

Whether and when a parent is admitted, which determines $A$, is itself governed by a probability of hospitalisation and a delay from symptom onset to admission. Both are permitted to vary over calendar time, representing improving access to and timeliness of care as the outbreak response matures, and are described in detail in **Step 5**. Each secondary infection is then assigned a host class: the newly infected individual is a HCW with a probability $q_{j,s_k}$ that depends on the setting of transmission $s_k$ and the class $j$ of the parent:

$$\Pr(\text{offspring } k \text{ is a healthcare worker}) = q_{j,\,s_k}, \qquad q_{j,\text{hospital}} > q_{j,\text{community}},$$

so that HCWs make up a (relatively speaking) larger share of those infected in the hospital than in the community, capturing their elevated occupational exposure. The assigned setting and class together determine which interventions can act on each infection, as described next.

**Step 3: Reducing transmission in health-care settings: ETU isolation and PPE/IPC:** Two NPIs reduce transmission in the hospital setting: the isolation of admitted cases within specialised health-care facilities, and the use of PPE and IPC by HCWs. For each, we distinguish its coverage, the time-varying probability that the measure reaches a given case, from its efficacy, the fixed proportional reduction in transmission it achieves once in place. Coverage is the lever that strengthens as the outbreak response matures; efficacy reflects the intrinsic effectiveness of the measure.

The first intervention reduces transmission from cases once they have been admitted and isolated, with the size of the reduction depending on the type of facility in which a case is managed. Cases admitted to a dedicated ETU, with isolation and rigorous infection control, are subject to a high transmission-reduction efficacy $e_{ETU}$, whereas those managed in general health-care facilities are subject to a substantially lower efficacy $e_{gen}$. We represent this distinction explicitly through the proportion of admitted cases managed in an ETU, $\pi(t)$, which serves as the coverage for this intervention and rises over calendar time as ETU capacity is established and operationalised. The effective post-admission transmission-reduction efficacy is then the coverage-weighted average of the two facility types:

$$q(t) = \pi(t)\, e_{\text{ETU}} + \big(1 - \pi(t)\big)\, e_{\text{gen}},$$

so that a transmission from an admitted, isolated source (all hospital transmission from a general-population source, and post-admission hospital transmission from a HCW source) is retained with probability

$$\Pr(\text{retained} \mid \text{isolation}) = 1 - q(t).$$

The second intervention, PPE and IPC, reduces transmission involving HCWs in the hospital, either by protecting a HCW exposed while caring for an infectious patient or by reducing onward transmission between workers that involves an infected HCW who continues working before their own admission (again, we note this is included for the sake of completeness but this fraction is very small in practice). Its coverage $c(t)$, the probability that the relevant health-care worker is adequately protected, likewise rises over time as supplies and practice improve, while its efficacy $e_{PPE}$ (the efficacy of the PPE when available and being worn correctly) is fixed. A protected hospital interaction is retained with probability:

$$\Pr(\text{retained} \mid \text{PPE}) = 1 - c(t)\, e_{\text{PPE}}.$$

These interventions act as independent layers. Where more than one applies to the same transmission event, for instance a post-admission hospital infection, which is subject to both isolation (of the infected individual) and PPE (worn by the HCW treating the infected individual), the transmission is retained only if it is not removed at each layer, so that its overall retention probability is the product of the relevant layer-specific probabilities.

**Step 4: Funeral transmission and SDBs:** Transmission at funerals is a defining feature of ebolavirus epidemiology[6,9], in which contact with the body of a person who has died of the disease, during traditional preparation and burial, can generate large clusters of secondary infections. We model this route separately, and only for cases who die: survivors generate no funeral transmission. When a case dies, the number of secondary infections arising from their funeral is drawn from a separate negative binomial distribution:

$$N_F \sim \text{NegBin}(\text{mean} = R_F(t_{\text{death}}),\ \text{size} = k_F)\,,$$

where $R_F$ is the funeral transmissibility, evaluated at the time of death so that it too may change as the outbreak progresses, and $k_F$ captures the marked overdispersion of funeral-associated transmission, through which a single unsafe burial can seed many infections. Because a funeral is a discrete event occurring shortly after death, the timing of each funeral infection is set by a short Gamma-distributed delay applied to the time of death,

$$\tau_F \sim \mathrm{Gamma}(\alpha_F, \beta_F),$$

so that funeral infections are concentrated around the burial rather than spread across an infectious period.

SDBs act as a NPIoperating on this exposure and transmission pathway, and as elsewhere we distinguish coverage from efficacy. Coverage is defined as the probability that a given death leads to an unsafe burial $u(t)$, which declines over calendar time as SDB teams are established, and which we allow to differ by the setting in which the death occurred (community or hospital) and the class of the deceased, since deaths in care are more readily managed with safe procedures than deaths in the community. Whether a particular funeral is safe or unsafe is determined by a single draw with this probability. An unsafe funeral generates the full complement of secondary infections drawn above, whereas at a safe funeral each candidate infection is retained only with probability $(1-s)$, where the fixed efficacy $s$ is the proportional reduction in transmission achieved by a safe and dignified burial. Marginalising over burial safety, a candidate funeral infection is therefore retained with probability

$$\Pr(\text{retained}) = 1 - s\left[1 - u(t_{\text{death}})\right],$$

These retained funeral infections enter the transmission tree and, like all secondary infections, are assigned a host class (Step 2), though funeral transmission predominantly involves the general population.

**Step 5: Completing the clinical course and natural history of each generated infection:** Each case arising as a result of a retained secondary infection is then carried through its own clinical course, which determines whether and when it is hospitalised, whether it dies, and, for those who die, the safety of the ensuing burial. These attributes in turn govern that case's onward transmission when it is later selected as a parent. We first determine symptomatic status with probability $p_{symp}$, and assign a symptom-onset time by adding a drawn incubation period to the infection time. Only symptomatic cases can progress to hospitalisation or death; asymptomatic infections recover without doing either (though in these analyses we assume all ebolavirus infections lead to symptoms). For each symptomatic case we next determine potential hospitalisation. Admission is drawn with a probability $p_{hosp,j}(t)$ that is specific to the case's class $j$ (general population or HCW) and evaluated at the time of symptom onset, so that it can rise as access to care improves over the course of the outbreak. The interval from symptom onset to admission is drawn from a delay distribution and scaled by a time-varying factor, again evaluated at symptom onset, allowing the timeliness of care to change as the response matures. We call this hospitalisation "potential" because whether it is actually realised depends on the timing of the case's outcome i.e. an individual can only be successfully hospitalised if (and only if) they do not die prior to hospitalisation occurring.

In order to reflect this, we first assign a provisional community course: the case would die with probability $p_{death,comm}$ and otherwise recover, with the time of this provisional outcome drawn from the corresponding onset-to-death or onset-to-recovery distribution. A potentially hospitalised case is admitted only if its admission time precedes this provisional outcome, capturing the fact that a case which dies or recovers at home before reaching care is never hospitalised. Among those admitted, a case whose provisional course was recovery also recovers in care, whereas a case whose provisional course was death is given a fresh survival draw, dying in care with probability $p_{death,hosp}/p_{death,comm}$ and surviving otherwise.

$$\Pr(\text{die} \mid \text{symptomatic, community}) = p^{\text{comm}}_{\text{death}}, \qquad \Pr(\text{die} \mid \text{admitted}) = p^{\text{hosp}}_{\text{death}} \le p^{\text{comm}}_{\text{death}}.$$

This second chance yields a realised in-care case fatality risk of exactly $p_{death,hosp}$, which we assume to be smaller than the community case fatality risk $p_{death,comm}$, so that timely admission to care confers a survival benefit. The time of the realised outcome is measured from admission for those who are hospitalised and from symptom onset otherwise. For every case who dies we assign the safety of their burial with the setting- and class-specific probability of an unsafe burial introduced in Step 4, evaluated at the time of death; this determines whether the death will later generate funeral transmission when the case is processed as a parent. The completed infection is then added to the transmission tree, and control returns to Step 1, where the next earliest unprocessed infection is selected, until no further infections remain and the outbreak has ended.

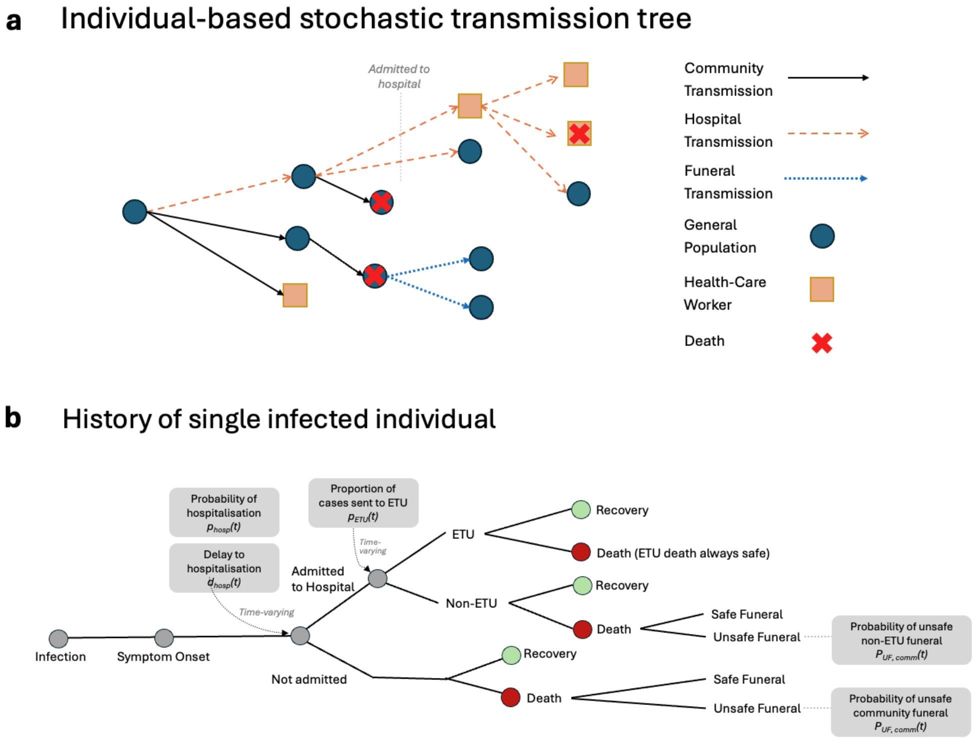


**Figure S1. Schematic of the stochastic branching-process model.**
**a)** Individual-based stochastic transmission tree. The model propagates infections through calendar time, with each case generating secondary infections in community, health-care, or funeral settings. Infections are assigned to either the general population or health-care workers, and fatal infections can subsequently generate funeral-associated transmission. **b)** Clinical and response pathway for a single infected individual. Each infection progresses from infection to symptom onset and then either to hospital admission or non-admission, followed by recovery or death. Hospitalised cases can be managed in an Ebola treatment unit (ETU) or non-ETU setting, and deaths are followed by safe or unsafe funerals depending on setting-specific, time-varying probabilities. Time-varying response parameters include the probability of hospitalisation, delay to hospitalisation, proportion of admitted cases managed in an ETU, and probabilities of unsafe funeral following community or non-ETU hospital death. ETU=Ebola treatment unit; HCW=health-care worker.

#### 1.3 Modelling the Impact of Antivirals on HCW Disease Burden

Orally available antivirals have shown protective efficacy against filoviruses when administered after exposure[7,8]. We model their use as post-exposure prophylaxis for HCW, the group at the most concentrated occupational risk during ebolavirus outbreaks and whose infection both removes scarce clinical capacity and seeds further nosocomial transmission. In contrast to the NPIs described above, which reduce the probability that an exposure results in transmission, antivirals act pharmacologically after exposure has occurred, preventing an exposed individual from going on to develop infection (and subsequent morbidity and mortality). By default the drug is offered to health-care workers exposed in the hospital setting, although our framework is flexible and can enable targeting of other groups and settings as applicable. For each eligible exposure, three quantities determine whether antivirals prevent resulting infection. The drug is received with probability $c_{OBV}(t)$, its coverage, which may rise over calendar time as supply and delivery improve; a received course is adhered to sufficiently for protection with probability $a_{OBV}$, its adherence. Conditional on receipt and adherence, the drug prevents infection with an efficacy $E$. The probability that antivirals prevent a given eligible infection is therefore:

$$\Pr(\text{infection prevented}) = c_{\text{OBV}}(t)\, a_{\text{OBV}}\, E,$$

so that the value of prophylaxis depends jointly on how many exposed HCWs receive and adhere to it. Infections that are prevented are removed before they can progress to disease, death, or onward transmission. We distinguish two policies for deciding which exposed HCWs are offered antivirals, defined by where they sit relative to the protective effect of PPE and ETU isolation. Under the first policy, antivirals are offered to every HCW who experiences an infectious exposure in the hospital, before accounting for whether PPE or isolation in an ETU would, on its own, have prevented the resulting infection; this policy sits upstream of those interventions and treats all exposed HCWs. Under the second policy, antivirals are offered only to the subset of exposed HCWs whose exposure is not averted by PPE or ETU isolation, that is, those for whom these protections have failed and who would otherwise become infected; this policy sits downstream of the two interventions and targets only these high-risk, protection-failure exposures. Because only infections that would otherwise occur can be prevented, the two policies avert the same infections, and so achieve the same reduction in health-care workers disease burden; they differ in the number of courses that must be delivered to do so, with the first treating many workers who were already protected and the second confining treatment to those who were not; though the second assumes perfect information on who would go on to be infected. These two policies therefore represent an upper and lower bound on the number of doses required. The implications of this for the number of doses required, and hence for stockpiling and the efficiency with which a fixed supply of drug is used, are described in detail in a subsequent section where we outline the different simulation analyses carried out. Across either policy, antivirals reduce the HCW disease burden by preventing a fraction of the infections that PPE and ETU isolation do not. The model records the infections prevented and deaths averted among HCWs, allowing the impact of antivirals, and their dependence on coverage, adherence, and the timeliness of dosing, to be quantified directly.

#### 1.4 Model Calibration to Past Outbreak Dynamics

Historical ebolavirus outbreaks provide an empirical basis for ensuring that simulated epidemics reproduce the scale, timing, and health-care workers burden observed in previous events. We therefore used sequential Approximate Bayesian Computation (ABC), implemented in the R package *easyABC*, to calibrate the branching-process model to two large ebolavirus outbreaks: the 2013-2016 epidemic in West Africa and the 2018-2020 outbreak North Kivu and Ituri in eastern Democratic Republic of the Congo (DRC). These outbreaks provide two different calibration settings. The West Africa epidemic was much larger, with a higher absolute mortality burden and a large number of health-care workers deaths. The North Kivu and Ituri outbreak was smaller but occurred in a setting affected by insecurity and conflict that led to sustained transmission and a prolonged outbreak. We did not explicitly model ring vaccination. This reflects the primary policy question addressed here: the role of antiviral PEP in outbreaks where species-specific vaccines are unavailable, delayed, or not readily deployable, as in Bundibugyo virus disease. This choice affects interpretation of the DRC-like archetype, because rVSV-ZEBOV-GP vaccination was deployed during the 2018–2020 North Kivu and Ituri outbreak. The DRC-like archetype should therefore be interpreted as a calibrated conflict-disrupted response setting, not as a mechanistic reconstruction of vaccination, NPIs, and case management as separable interventions. The fitted parameters reproduce aggregate realised outbreak dynamics, including the net effect of the response environment observed during the outbreak.

For each outbreak, we fitted five parameters governing the main transmission and exposure processes in the model: the basic reproduction number (R0), the proportion of overall transmission attributable to funerals, separate scalars controlling PPE efficacy and ETU isolation efficacy, and a scalar governing the intensity of health-care setting exposure experienced by HCWs. These parameters were fitted to a set of epidemic-scale summaries capturing the size, timing, and health-care workers burden of each outbreak: the proportion of stochastic replicates in which the epidemic became established (defined as exceeding 100 cumulative deaths), total mortality, mortality among HCWs, the fraction of deaths occurring among HCWs, the height of the peak in weekly Ebola virus disease mortality (i.e. the maximum weekly incidence of Ebola virus disease mortality observed), and the central-90% death interval, defined as the number of days separating the 5th and 95th percentiles of the death-date distribution (equivalently, the span covering the central 90% of cumulative deaths). The three count summaries (total deaths, HCW deaths, and peak weekly mortality) were matched on the log scale in order to capture the relative, fold-differences in modelled and empirical estimates. We used the central-90% death interval in preference to a first-to-last-death duration because it is robust to the early seeding period and to the late sporadic chains and terminal low-incidence tails that make a full-span duration noisy and sensitive to isolated late events. Because detailed transmission-chain data are sparse, incomplete, and variable in quality across these outbreaks, calibration was performed against these summary features of the main epidemic trajectory rather than against individual observed infections or transmission links. The resulting outbreak-specific parameter posterior distributions were used to define two representative ebolavirus epidemic “archetypes,” calibrated to realised historical outbreak dynamics rather than to the separate mechanistic effects of each intervention, and spanning a plausible range of severe historical outbreak dynamics, which then served as reference settings for evaluating the potential impact of antivirals. This allowed the drug’s impact to be assessed within simulated outbreaks that were consistent with the scale, temporal structure, funeral contribution, intervention context, and HCW burden observed in previous ebolavirus epidemics.

For each outbreak, we first constructed an outbreak-specific baseline parameterization of the model. This included fixed natural-history parameters governing incubation period, symptom onset, hospitalisation, recovery, and death, alongside time-varying inputs describing how the outbreak response changed over calendar time. These time-varying inputs included the probability that symptomatic cases were hospitalized, the delay from symptom onset to hospitalisation, the proportion of hospitalised cases managed in ETUs, the coverage of PPE and IPC among HCWs, and the probability that deaths were followed by unsafe funerals. These quantities were estimated separately from literature-review and outbreak-response data, as described below (and see **Figure S2**), and were then supplied to the ABC calibration as fixed functions of time rather than fitted parameters. For each proposed ABC parameter set, these fixed and time-varying inputs were combined with the proposed values of R0, the proportion of transmission attributable to funerals, PPE efficacy, ETU isolation efficacy, and health-care setting exposure among health-care workers to generate a complete set of inputs for the branching-process model. Multiple stochastic outbreaks were then simulated for each proposed parameter set, retaining those that became established (≥100 cumulative deaths), and the retained epidemics were summarized using the same summaries used to define the observed calibration targets: total mortality, health-care workers mortality, the HCW death fraction, peak weekly mortality, and the central-90% death interval.

For the West Africa outbreak calibration, the fitted targets were 11,325 total deaths[10] and 513 HCW deaths, giving a HCW death fraction of 0.045[11]. The peak-week death count was 599 and the duration of the central90% of deaths I was~274 days, calculated from the reported death time series rather than taken from a published figure [12]. For the North Kivu and Ituri calibration, the corresponding targets were 2,299 total deaths and 79 HCW deaths giving a HCW death fraction of 0.034[13,14], with a peak-week count of 95 deaths and a central-90% death duration of ~378 days, derived from the reported time series[15]. The central-90% death interval was computed identically for observed and simulated outbreaks, as the span between the days on which cumulative deaths reached 5% and 95% of the outbreak total; no separate delimitation of a "main epidemic phase" was therefore required, and the timing target is insensitive to isolated late chains of transmission.

Calibration was implemented using the Del Moral sequential ABC algorithm in *EasyABC*. For both outbreak archetypes, we assigned uniform prior distributions to the five fitted parameters: R0 ~ Uniform(1.15, 1.65), the proportion of transmission attributable to funerals ~ Uniform(0.10, 0.40), PPE efficacy ~ Uniform(0.3, 0.9), ETU efficacy ~ Uniform(0.6, 0.9) and the health-care workers risk scalar ~ Uniform(1.0, 3.0). The health-care workers risk scalar multiplies a baseline probability of 0.25 that a hospital transmission event infected a health-care worker, rather than a member of the general population. Each model simulation was initialized with 25 seeding infections. Peak weekly mortality was calculated using 7-day death-incidence bins. Simulated epidemics were considered to have taken off if they generated at least 100 deaths; take-off probability was calculated across stochastic replicates, while the remaining calibration summaries were calculated among replicates that reached this threshold. Each calibration was run using 100 stochastic replicates per ABC particle and 500 ABC particles total, with a target tolerance of 0.5, and yielded a posterior sample of the five fitted parameters. These posterior samples, comprising R0, the proportion of transmission attributable to funerals, PPE and ETU efficacy, and the HCW risk scalar, were then used to generate the baseline outbreak trajectories against which antiviral scenarios were compared. For each posterior draw, we simulated a no-antiviral scenario, representing the fitted historical epidemic archetype as it would occur in the absence of antivirals (as was the case during the historic epidemics), and compared this with otherwise matched scenarios in which PEP was available. This paired counterfactual structure allowed the estimated impact of antivirals to be calculated as the difference between matched simulations with and without PEP, while propagating uncertainty in the fitted historical outbreak dynamics into estimates of infections prevented and HCW deaths averted.

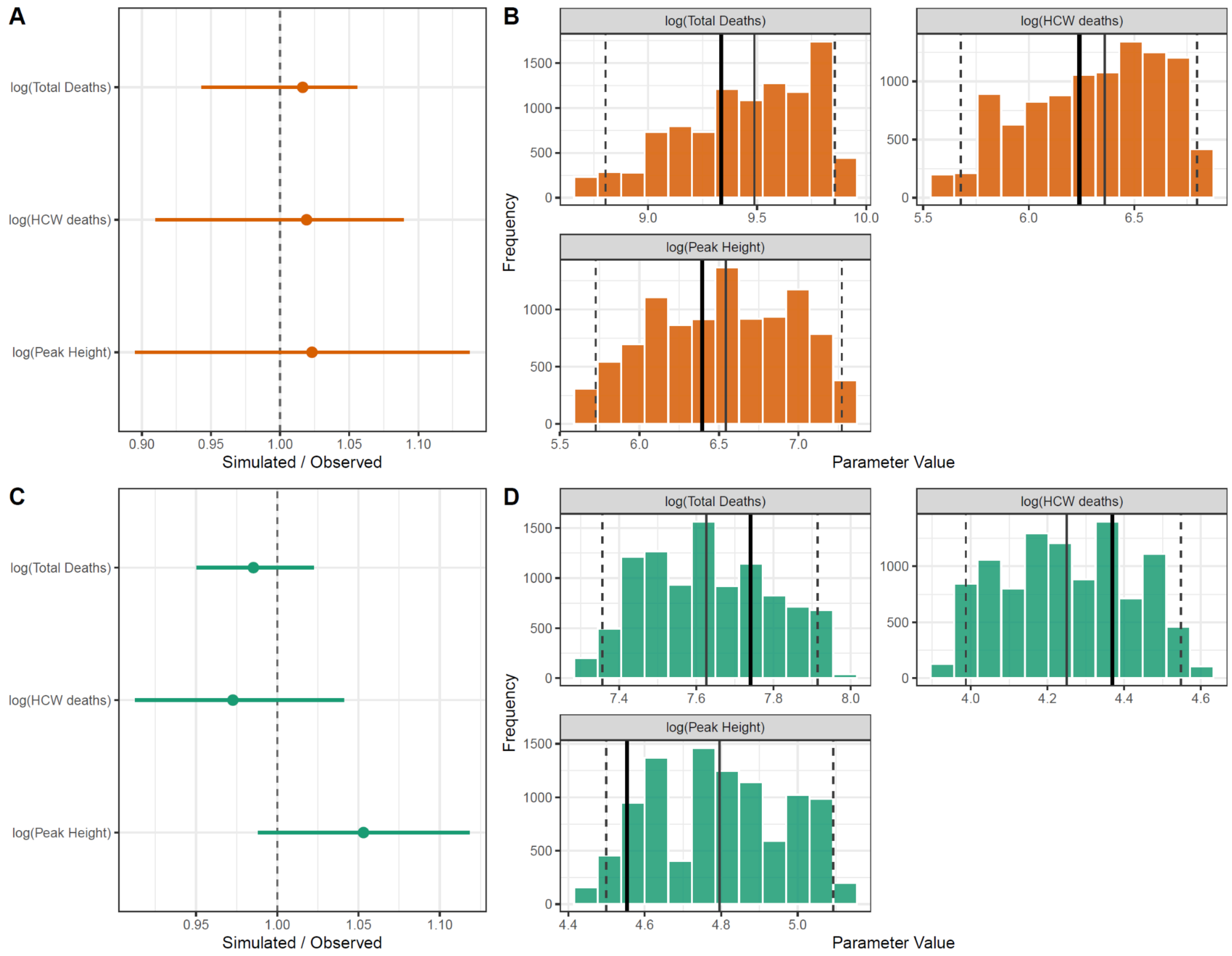


**Figure S2: Posterior predictive fit to historical outbreak calibration targets.** Posterior predictive checks for the sequential approximate Bayesian computation calibration of the branching-process model to the "West Africa-like" and "DRC-like" outbreak archetypes. **a)** Ratio of simulated to observed calibration summaries for the West Africa-like archetype. Points show posterior predictive medians and horizontal lines show 95% credible intervals for each fitted summary statistic. The dashed vertical line indicates perfect agreement between simulated and observed values. **b)** Posterior predictive distributions of the fitted West Africa calibration summaries on the log scale: total deaths, health-care workers deaths, and peak weekly deaths. Histograms show the distribution of simulated summaries across posterior-predictive draws; thick vertical lines indicate observed calibration targets, thin vertical lines indicate posterior predictive medians, and dashed vertical lines indicate 95% credible intervals. **c)** Ratio of simulated to observed calibration summaries for the DRC-like archetype, plotted as in a. **d)** Posterior predictive distributions of fitted calibration summaries for the DRC-like archetype, plotted as in b. Orange indicates the West Africa-like archetype and green indicates the DRC-like archetype. HCW=health-care worker; DRC= Democratic Republic of Congo

#### 2. Supplementary Methods: Data Sources and Model Parameterisation

We parameterised the branching-process model using a targeted review of published outbreak investigations, epidemiological analyses, mathematical modelling studies, and operational reports from WHO, US CDC, national ministries of health, UNMEER, and response partners. Sources were prioritised when they reported values relevant to Ebola virus disease natural history, transmission heterogeneity, health-care workers infection risk, clinical outcomes, hospitalisation, ETU care, IPC/PPE, and SDBs. Where multiple estimates were available, we preferentially used values from the outbreak setting most closely aligned with the corresponding archetype, distinguishing between the 2013–2016 West Africa epidemic and the 2018–2020 North Kivu/Ituri outbreak in eastern DRC. Fixed model inputs are summarised in **Table S1**, while source-reported values used to inform the time-varying response-parameter trajectories are summarised in **Table S2**.

##### 2.1 Fixed, non-time-varying model parameters

Static model parameters included fixed intervention efficacies, offspring overdispersion, generation-time distributions, natural-history delays, case fatality risks and HCW infection-allocation probabilities. These parameters were treated as fixed inputs within each outbreak archetype, with separate "West Africa-like" and "DRC-like" values used where setting-specific evidence supported different parameterisation. 'Value used in simulations' denotes the parameter values used in the primary analyses, while 'Range across collated literature' gives the minimum to maximum values evidenced from the targeted literature review. See **Table S1** for the full list of parameters.

##### 2.2 Source values for time-varying response parameters

For response parameters that changed over the course of an outbreak, we collated source-reported values from outbreak investigations, modelling studies, WHO situation reports, response plans, operational datasets, and safe-burial performance analyses. These values were not used directly as stepwise model inputs. Instead, they were used to inform the fitted outbreak-specific response trajectories described in Section 2.3. **Table S2** summarises the source-reported values used to inform each time-varying parameter trajectory, including values for the West Africa-like and DRC-like outbreak archetypes where different evidence sources informed the corresponding curve.

**Table S1. Static model parameters used in the  branching-process simulations.**
Fixed parameter values used in the branching-process model, stratified by outbreak archetype where values differed between the “West Africa-like” and “DRC-like” settings. Central values denote the baseline values used in the primary analyses, while ranges denote literature-supported minima and maxima. Time-varying response parameters are reported separately in Table S2.

| Model component | Description | Archetype | Value used in simulations | Range across collated literature | Units | Sources reviewed |
|---|---|---|---|---|---|---|
| **NPI Efficacy** | | | | | | |
| ETU efficacy | Fixed proportional reduction in onward hospital transmission for cases managed in an ETU | Both | Estimated as part of calibration | NA | Proportion | NA |
| PPE/IPC efficacy | Fixed proportional reduction in health-care workers infection risk when PPE and IPC are available and correctly implemented. Coverage varies over time through the IPC/PPE response curve (Table S2). | Both | Estimated as part of calibration | NA | Proportion | NA |
| Non-ETU hospital efficacy | Fixed proportional reduction in hospital transmission for admitted cases managed outside dedicated ETU-level isolation. | West Africa | 0.30 | 0.05–0.61 | Proportion | West Africa:[9,16–19]; DRC: [5,20–22] |
| | | DRC | 0.30 | 0.05–0.50 | | |
| Safe and dignified burial efficacy | Fixed proportional reduction in funeral-associated transmission when a burial is managed safely. Implementation and coverage are represented separately through the time-varying unsafe-funeral probabilities in Table S2. | Both | 0.80 | 0.33-1.00 | Proportion | [23–25] |
| **Transmission Heterogeneity/ Generation-Time** | | | | | | |
| Offspring distribution overdispersion | Negative-binomial dispersion parameter for general-population parent offspring. Assumed equal for general population and HCWs. | West Africa | 0.18 | 0.03–0.39 | Dimensionless | West Africa: [9,26–29]; DRC:[20] |
| | | DRC | 0.27 | 0.20–0.33 | | |
| Offspring distribution mean | Mean number of secondary infections generated by non-funeral transmission before allocation across host class and setting. | Both | Estimated as part of calibration | NA | Dimensionless | NA |
| Funeral offspring distribution overdispersion | Negative-binomial dispersion parameter for funeral-associated offspring. | Both | 0.30 | 0.03–0.33 | Dimensionless | [9,24,26–29] |
| Funeral offspring distribution mean | Mean number of funeral-associated secondary infections. | Both | Estimated as part of calibration | NA | Dimensionless | NA |
| Generation-time distribution general population and HCW | Gamma-distributed generation-time kernel, parameterised by shape and rate. Values used in simulations give a mean generation time of 15.4 days. | Both | Shape: 2.50<br>Rate: 0.16 | Shape: 2.50-5.33<br>Rate: 0.16-0.44 | Shape: Dimensionless<br>Rate: $day^{-1}$ | [6,28,30] |

| | | | | | | |
|---|---|---|---|---|---|---|
| Generation-time distribution funeral | Gamma-distributed generation-time kernel, parameterised by shape and rate. Values used in simulations give a mean generation time of 2.0 days. | Both | Shape: 20.00<br>Rate: 10.00 | NA | Shape: Dimensionless<br>Rate: $day^{-1}$ | Modelling Assumption |
| **Natural History** | | | | | | |
| Incubation period distribution | Gamma distribution parameterised by mean delay from infection to symptom onset and associated standard deviation | Both | 8.50<br>Assumed SD: 4.5 | 7.70–11.40 | Days | [6,28,30] |
| Onset to death | Gamma distribution parameterised by mean delay from symptom onset to death among fatal cases and associated standard deviation | Both | 9.30<br>Assumed SD: 3.0 | 8.50–10.10 | Days | [6,30,31] |
| Onset to recovery | Gamma distribution parameterised by mean delay from symptom onset to recovery among non-fatal cases (not admitted to care) and associated standard deviation. | Both | 13.00<br>Assumed SD: 4.0 | 10.40–15.70 | Days | [6,30,31] |
| Symptomatic infection probability | Probability that infection becomes symptomatic. | Both | 1 | 1 | Probability | Modelling Assumption |
| Community death probability | Probability of death for symptomatic cases whose clinical course is completed in the community. | West Africa | 0.71 | 0.60–0.74 | Probability | West Africa:[6,32,33]; DRC:[13,22,34] |
| | | DRC | 0.70 | 0.63–0.76 | | |
| Hospital death probability | Probability of death for symptomatic cases admitted to care. | West Africa | 0.34 | 0.20–0.50 | Probability | West Africa:[6,9,32]. DRC:[13,22,34] |
| | | DRC | 0.49 | 0.24–0.59 | | |
| **Hospitalisation-related** | | | | | | |
| Delay to hospitalisation | Gamma distribution parameterised by mean delay from symptom onset to hospital admission and associated standard deviation (before applying time-varying response scaling). | West Africa | 4.50<br>Assumed SD: 0.35 | 2.30–5.00 | Days | West Africa:[6,9,28,35] ;DRC:[5,20–22] |
| | | DRC | 3.50<br>Assumed SD: 0.35 | 2.80–4.10 | | |
| Hospitalisation to death | Gamma distribution parameterised by mean delay from hospital admission to death among hospitalised fatal cases and associated standard deviation | West Africa | 4.50<br>Assumed SD: 2.0 | 3.50–7.80 | Days | West Africa:[28,30,35] ;DRC:[21,22,30] |
| | | DRC | 4.50<br>Assumed SD: 2.0 | 4.40–7.30 | | |
| Hospitalisation to recovery | Gamma distribution parameterised by mean delay from hospital admission to recovery among hospitalised non-fatal cases and associated standard deviation | West Africa | 8.00<br>Assumed SD: 2.5 | 5.40–13.40 | Days | West Africa:[28,30,35] ;DRC:[21,22,30] |
| | | DRC | 8.00<br>Assumed SD: 2.5 | 6.30–12.90 | | |
| **HCW Exposure/Allocation** | | | | | | |

| | | | | | | |
|---|---|---|---|---|---|---|
| HCW infection allocation | Probability that a community transmission event from a general-population parent infects a health-care worker. | West Africa | 0.005 | 0.01–0.08 | Probability | West Africa:[9,11,36–38] ;DRC:[5,13,38] |
| | | DRC | 0.005 | 0.02–0.08 | | |
| HCW infection allocation | Probability that a hospital transmission event from a general-population parent infects a health-care worker. This value is estimated during calibration but set to a baseline value of 0.25. | West Africa | 0.25 | 0.08–0.49 | Probability | West Africa:[5,9,11,28] DRC:[5,9,38] |
| | | DRC | 0.25 | 0.16–0.49 | | |
| HCW infection allocation | Probability that a community transmission event from a HCW parent infects another HCW. | Both | 0.02 | 0.01–0.08 | Probability | [9,11,36–38] |
| HCW infection allocation | Probability that a hospital transmission event from a HCW parent infects another health-care worker. This value is estimated during calibration but set to a baseline value of 0.25 | West Africa | 0.25 | 0.08–0.49 | Probability | West Africa:[5,9,11,28] ;DRC:[5,9,38] |
| | | DRC | 0.25 | 0.16–0.49 | | |
| HCW hospital exposure allocation | Probability that a pre-admission transmission event from an infected HCWr occurs in a health-care setting. | West Africa | 0.49 | 0.08–0.72 | Probability | West Africa:[5,9,11,28] ;DRC:[5,9,38] |
| | | DRC | 0.50 | 0.17–0.72 | | |
| Funeral HCW allocation | Probability that a funeral-associated transmission event from a HCW descendant infects a HCW. | Both | 0.02 | 0.01–0.12 | Probability | [9,23,24,39,40] |
| Funeral HCW allocation | Probability that a funeral-associated transmission event from a general-population decedent infects a HCW. | Both | 0.005 | 0.01–0.12 | Probability | [9,23,24,39,40] |

**Table S2. Literature-derived values used to inform time-varying response-parameter trajectories.** Time-varying response parameters used in the branching-process model, with source-reported values used to inform the fitted outbreak-specific response trajectories. Rows are grouped by model parameter, with separate entries for the West Africa-like and DRC-like outbreak archetypes where different data sources informed the corresponding trajectory. Source-reported values are given by relative outbreak day and parameter value; where multiple estimates were available for the same time point, values are shown as ranges. These values correspond to the plotted points in the fitted response-parameter figures. The resulting daily trajectories were estimated separately from the ABC calibration and supplied to the branching-process model as fixed outbreak-specific input functions.

| Time-varying parameter | Definition | Direction | Archetype | Source-reported value(s) used to inform curve | Units | Source(s) |
|---|---|---|---|---|---|---|
| d_hosp(t) | Delay from symptom onset to hospitalisation/isolation; applied as the time-varying mean delay. | Decreases with response maturation | West Africa | relative day 31: 5; relative day 107: 4.6 | days | 6 |
| | | | West Africa | relative day 39: 5.3 | days | 33 |
| | | | West Africa | relative day 84: 4.4; relative day 267: 3; relative day 357: 2.3 | days | 35 |
| | | | West Africa | relative day 169: 4; relative day 283: 3.6 | days | 41 |
| | | | West Africa | relative day 176: 4.5 | days | 28 |
| | | | West Africa | relative day 264: 4.1 | days | 42 |
| | | | DRC | relative day 0: 4.8 | days | 43,44 |
| | | | DRC | relative day 44: 2–4 | days | 21 |
| | | | DRC | relative day 212: 4.2 | days | 44 |
| p_hosp(t) | Probability that a symptomatic infected individual is hospitalised/isolated. | Increases with response maturation | West Africa | relative day 0: 0.35 | proportion | 33 |
| | | | West Africa | relative day 196: 0.51 | proportion | 45 |
| | | | West Africa | relative day 203: 0.55 | proportion | 19 |
| | | | West Africa | relative day 245: 0.85 | proportion | 46 |
| | | | West Africa | relative day 248: 0.7 | proportion | 47 |
| | | | West Africa | relative day 255: 0.7 | proportion | 48 |
| | | | DRC | relative day 7: 0.364 | proportion | 43 |
| | | | DRC | relative day 168: 0.6 | proportion | 21 |
| | | | DRC | relative day 212: 0.5 | proportion | 44 |

| | | | | | | |
|---|---|---|---|---|---|---|
| p_ETU(t) | Proportion of hospitalised cases managed in ETU care. | Increases with response maturation | West Africa | relative day 0: 0 | proportion | 35,49 |
| | | | West Africa | relative day 195: 0.212 | proportion | 50 |
| | | | West Africa | relative day 203: 0.2–0.364 | proportion | 19 |
| | | | West Africa | relative day 276: 0.741 | proportion | 51 |
| | | | DRC | relative day 0: 0.35 | proportion | 43 |
| | | | DRC | relative day 44: 0.846 | proportion | 21 |
| | | | DRC | relative day 212: 0.55 | proportion | 44 |
| I_IPC(t) | Latent IPC/response-maturity index used as the time-varying IPC/PPE input. | Increases with response maturation | West Africa | relative day 0: 0.05 | index (0–1) | 52 |
| | | | West Africa | relative day 194: 0.1 | index (0–1) | 53 |
| | | | West Africa | relative day 210: 0.3 | index (0–1) | 45,54 |
| | | | West Africa | relative day 313: 0.8 | index (0–1) | 55 |
| | | | DRC | relative day 226: 0.2 | index (0–1) | 56 |
| | | | DRC | relative day 300: 0.65 | index (0–1) | 44,57 |
| | | | DRC | relative day 457: 0.5 | index (0–1) | 57 |
| p_UF,comm(t)[†] | Probability that a community death results in an unsafe funeral. | Decreases with response maturation | West Africa | relative day 0: 1 | proportion | 58,59 |
| | | | West Africa | relative day 176: 0.119 | proportion | 28 |
| | | | West Africa | relative day 255: 0.3 | proportion | 48 |
| | | | West Africa | relative day 262: 0.23 | proportion | 60 |
| | | | West Africa | relative day 301: 0 | proportion | 61 |
| | | | DRC | relative day 0: 0.6 | proportion | 40,62 |
| | | | DRC | relative day 273: 0.39 | proportion | 40 |
| | | | DRC | relative day 333: 0.2 | proportion | 44 |
| | | | DRC | relative day 333: 0.15 | proportion | 62 |
| p_UF,hosp(t) | Probability that a non-ETU hospital death results in an unsafe funeral. | Decreases with response maturation | West Africa | relative day 176: 0.02 | proportion | 28,45 |

| | | | | | | |
|---|---|---|---|---|---|---|
| | | | West Africa | relative day 245: 0 | proportion | 46 |
| | | | DRC | relative day 0: 0.01 | proportion | 40,43 |
| | | | DRC | relative day 333: 0 | proportion | 40,43 |

*† For the "DRC-like" archetype, p_UF,comm(t) was directly informed by the time-series safe and dignified burial performance data extracted from Warsame et al.[40]. These data were used as an empirical proxy to inform the broader DRC response-quality curves for other time-varying parameters.*

##### 2.3 Estimating time-varying response parameters

The dynamics of ebolavirus outbreaks are shaped by changes in the quality, reach, and timeliness of the public-health response over time. As an outbreak response matures, cases may be identified and admitted to care more rapidly, a greater proportion of hospitalized cases may be managed in ETUs, PPE and IPC may become more widely available to HCWs, and SDBI teams may become established. These processes therefore need to be represented dynamically in order to capture the dynamics observed during ebolavirus outbreaks.

To incorporate these processes into the branching-process model, we collated available literature-derived observations on response-related parameters and fitted a latent response-quality model that converts these sparse observations into smooth time-varying parameter curves. We used this framework to estimate time-varying trajectories for six response parameters used by the branching-process model: the delay from symptom onset to hospitalisation, denoted $delay_{hosp}(t)$; the probability that a symptomatic case is hospitalised, $p_{hosp}(t)$; the proportion of hospitalised cases managed in an ETU or , $p_{ETU}(t)$; a latent IPC or PPE index, $I_{IPC}(t)$; the probability that a community death is followed by an unsafe funeral, $p_{unsafe,comm}(t)$; and the probability that a hospital death is followed by an unsafe funeral, $p_{unsafe,hosp}(t)$. What follows is first a description of the overall approach to representing these time-varying parameters in our modelling framework, followed by a description of the fitting process used to infer them.

For each outbreak archetype, we represented response maturation through a latent response-quality function $Q_a(tau)$, where $a$ indexes the outbreak archetype and tau is normalised outbreak time. We define normalised time as:

$$\tau = \frac{t}{T_a},$$

where $t$ is time in days since the start of the response curve and $T_a$ is the archetype-specific response horizon. The response-quality metric is constrained to lie between 0 and 1, with $Q_a(tau) = 0$ representing the least mature response state and $Q_a(tau) = 1$ representing the most mature response state reached over the calibration window. This response-quality curve is scale-free. Each model parameter is then mapped onto this shared response scale using its own minimum and maximum values, so that parameters with different units can change coherently over time while preserving parameter-specific magnitudes.

For a response parameter j in archetype a, let $L_{a,j}$ and $U_{a,j}$ denote the minimum and maximum values for that parameter over the course of an outbreak, with $L_{a,j}$ < $U_{a,j}$. For notational convenience, we write $Q_{a,j}(tau)$ for the response-quality curve applied to parameter $j$. For parameters that increase as the response matures, such as hospitalisation probability, ETU use, and IPC/PPE quality, the parameter trajectory is:

$$\theta_{a,j}(\tau) = L_{a,j} + (U_{a,j} - L_{a,j})\, Q_{a,j}(\tau).$$

For parameters that decrease as the response matures, such as time to hospitalisation and unsafe-funeral probabilities, the parameter trajectory is:

$$\theta_{a,j}(\tau) = U_{a,j} - (U_{a,j} - L_{a,j})\, Q_{a,j}(\tau).$$

However, direct measurements of these response processes are sparse, reported irregularly, and available on different scales for different outbreaks. For the West Africa archetype, we therefore estimated the response-quality dynamics jointly from collated information on each of the six response-parameters from the literature, using a partially pooled random effects logistic model. In this model, we explicitly separate and delineate the shape of the response curve (i.e. the underlying latent $Q_{a,j}(tau)$), which is fitted in a partially pooled approach and thus enables sharing of information between parameters; and the magnitude and exact value of each specific parameter (e.g. $delay_{hosp}(t)$) and its associated lower and upper bounds. Specifically, the shape parameters, which determine when the response improves and how rapidly it improves, were partially pooled across the six parameters. By contrast, the parameter-specific minimum and maximum values were estimated independently for each response parameter, i.e. the probabilities for hospitalisation, ETU use, PPE/IPC, and unsafe funerals, and days for the delay from symptom onset to hospitalisation. In the DRC archetype, the same empirical response-quality curve, derived from safe and dignified burial performance and available at fine temporal resolution across the outbreak, was used across all response parameters, so only the parameter-specific minimum and maximum values were estimated.

**"West Africa" archetype fitting:** For the West Africa archetype, the response-quality curve for parameter $j$ was represented using a finite-window logistic function. We first defined the unscaled logistic curve:

$$q^{\mathrm{raw}}_{a,j}(\tau) = \frac{1}{1 + \exp\left[-k_{a,j}\,(\tau - \tau_{50,a,j})\right]}.$$

Here $\tau_{50,a,j}$ is the normalized time at which the response process reaches its midpoint, and $k_{a,j}$ controls the steepness of improvement. Because the response window is finite, we rescaled this logistic curve so that each response-quality curve runs from exactly 0 to 1 across the observed window, $\tau \in [0,1]$:

$$Q_{a,j}(\tau) = \frac{q^{\mathrm{raw}}_{a,j}(\tau) - q^{\mathrm{raw}}_{a,j}(0)}{q^{\mathrm{raw}}_{a,j}(1) - q^{\mathrm{raw}}_{a,j}(0)}.$$

This normalisation makes $Q_{a,j}(\tau)$ interpretable as the fraction of response maturation achieved within the observed outbreak window, independent of the absolute scale of the parameter it modifies. The resulting response-quality curve was then mapped onto each parameter through its independently estimated minimum and maximum values ($L_{a,j}$ and $U_{a,j}$) as described above.

Partial pooling was applied to the West Africa curve-shape parameters, reflecting the assumption that different components of the outbreak response, including hospitalisation, ETU use, PPE/IPC, and safe burials, tend to improve over broadly similar time scales, while still allowing each process to depart from the shared pattern. Specifically, the midpoint and steepness parameters were estimated using a hierarchical non-centered parameterisation. For the midpoint, we used:

$$\operatorname{logit}(\tau_{50,a,j}) = \mu_{\tau 50,a} + \sigma_{\tau 50,a} z^{(\tau 50)}_{a,j}. \text{ and } z^{(\tau 50)}_{a,j} \sim \mathrm{Normal}(0,1).$$

and for the steepness, we used:

$$\log(k_{a,j}) = \mu_{\log k,a} + \sigma_{\log k,a} z^{(k)}_{a,j}. \text{ and } z^{(k)}_{a,j} \sim \mathrm{Normal}(0,1).$$

The hyperparameters $\mu_{\tau 50,a}$ and $\mu_{\log k,a}$ describe the average timing and steepness of response maturation across the six response parameters, while $\sigma_{\tau 50,a}$ and $\sigma_{\log k,a}$ determine how strongly individual parameter curves can deviate from that shared pattern. We assigned weakly informative priors centered on response curves that improve over the observed outbreak window:

$$\mu_{\tau 50,a} \sim \text{Normal}(0, 1.25).; \ \sigma_{\tau 50,a} \sim \text{HalfNormal}(0, 0.50).;$$
$$\mu_{\log k,a} \sim \text{Normal}\left(\log(8), 0.80\right).; \ \sigma_{\log k,a} \sim \text{HalfNormal}(0, 0.40).$$

This structure allowed sparse observations for one response process to inform the timing and speed of response maturation for others, while preserving parameter-specific trajectories through independently estimated minimum and maximum values. The partial-pooling model therefore shares information only over the dimensionless curve-shape parameters. It does not shrink the absolute parameter ranges toward a common value, because the response parameters correspond to different epidemiological quantities and have different units.

**"DRC" archetype fitting:** For the DRC archetype, we used a different approach because the available data structure differed from West Africa. Literature-derived observations for the six response parameters were still sparse, but, unlike for West Africa, a complete time series was available for one major component of the outbreak response: safe and dignified burial performance during the North Kivu and Ituri outbreak. We used this time series as an empirical proxy for the latent response-quality curve, rather than estimating a new smooth logistic response curve. This allowed the DRC archetype to preserve the observed non-monotonic response trajectory (i.e. the deterioration in response coverage and quality caused by the conflict and instability-imposed disruptions to the response that occurred during the outbreak), including periods of improvement, interruption, and deterioration, rather than forcing response quality to increase smoothly over time (as was the case with the "West Africa" archetype).

To construct this empirical response-quality curve, safe and dignified burial records were restricted to North Kivu and Ituri and aggregated by epidemiological week and province. For each province-week, we calculated the proportion of burial alerts with a successful or not-needed response as:

$$s_{p,w} = \frac{n_{p,w}^{\text{success}} + n_{p,w}^{\text{notneeded}}}{n_{p,w}^{\text{success}} + n_{p,w}^{\text{notneeded}} + n_{p,w}^{\text{failure}}}.$$

Here, $n_{p,w}^{\text{success}}$ is the number of successful safe and dignified burial responses in province $p$ and week $w$, $n_{p,w}^{\text{notneeded}}$ is the number of burial alerts for which no safe and dignified burial response was required, and $n_{p,w}^{\text{failure}}$ is the number of failed responses. False alerts and records with unclear outcomes were excluded. Province-specific weekly success curves were then averaged and smoothed using a centered four-week rolling mean. Let $\tilde{s}(t)$ denote this smoothed burial-response success curve. We then defined the DRC response-quality curve as:

$$Q_{\text{DRC}}(t) = \frac{\tilde{s}(t)}{\max_t \tilde{s}(t)}.$$

This scaling sets the best observed burial-response performance to $Q_{\text{DRC}}(t) = 1$, while preserving subsequent declines if response performance deteriorates later in the outbreak. Because the safe and dignified burial time series provides information on the absolute probability of unsafe funerals, the community unsafe-funeral probability was constructed directly from the smoothed success curve $p_{\text{unsafe,comm}}(t) = 1 - \tilde{s}(t).$ For the other time-varying response parameters, the empirical DRC response-quality curve $Q_{\text{DRC}}(t)$ was used as the shared latent curve, and the parameter-specific minimum and maximum values ($L_{a,j}$ and $U_{a,j}$) were estimated. Thus, in contrast to the West Africa archetype, the DRC model did not estimate parameter-specific response-curve shapes and did not apply partial pooling over curve-shape parameters. The fitted quantities were the lower and upper values of each response parameter, which mapped the shared empirical response-quality curve onto the appropriate parameter scale.

We additionally modified the DRC response-quality curve to represent the period of conflict-related disruption during which safe and dignified burial performance was not considered a reliable proxy for overall response quality. During this interval, we assumed that response capability returned to the level observed at the beginning of the outbreak. Operationally, this was implemented by setting the empirical response-quality curve back to its initial value over the conflict-disruption window before re-scaling the curve. This creates a "conflict-disrupted" DRC archetype in which all time-varying response parameters temporarily return to their least mature response state, before recovering according to the observed post-disruption trajectory. The resulting curve was then used as the shared $Q_{\mathrm{DRC}}(t)$ trajectory for hospitalisation probability, hospitalisation delay, ETU use, PPE/IPC coverage, and unsafe-funeral probabilities in the branching-process model.

Posterior distributions were estimated using Hamiltonian Monte Carlo, implemented in Stan[63]. Each model was run using four chains, with 1,500 warm-up iterations and 1,500 post-warm-up sampling iterations per chain. We used target acceptance probabilities of 0.97-0.98 and a maximum tree depth of 13. For the West Africa archetype, posterior samples were generated for the partially pooled response-curve shape parameters, the resulting parameter-specific latent response-quality curves $Q_{a,j}(\tau)$, the independently estimated minimum and maximum values $L_{a,j}$ and $U_{a,j}$, and the time-varying response trajectories implied by these quantities. For the DRC archetype, the response-quality curve $Q_{\mathrm{DRC}}(t)$ was fixed using the empirical safe and dignified burial performance time series, or its conflict-disrupted variant, and the posterior distribution was therefore estimated over the parameter-specific minimum and maximum values and the resulting mapped trajectories. The posterior mean of each time-varying parameter trajectory was carried forward into the branching-process simulations, while 90% posterior credible intervals were retained for visualising uncertainty in the fitted response curves (see **Figure S2**). The output of this fitting process was a daily set of time-varying model inputs for each Ebola epidemic archetype, the posterior means of which were then treated as fixed outbreak-specific inputs in the ABC calibration described above, so that the ABC procedure estimated baseline transmissibility, funeral contribution, NPI efficacy, and health-care workers exposure conditional on the inferred response-maturation trajectory for each archetype.

##### 2.4 Derivation of the non-human primate (NHP)-informed delay-to-initiation efficacy curve

We derived a delay-to-initiation efficacy curve for antiviral PEP from reconstructed NHP Kaplan–Meier survival data. These data were presented at the Keystone Symposium "Predicting and Responding to Emerging Viral Infections", Geneva, Switzerland, Oct 16, 2025. Associated presentation materials were reproduced for our analyses with relevant permissions. Survival times were extracted from step changes in the Kaplan–Meier curves, with one record generated per animal including treatment arm, dosing time in days post-challenge (DPC), time to death or administrative censoring, and event status **(Table S3).**

**Table S3.** Reconstructed NHP survival data used to fit the delay-to-initiation efficacy curve.

| Arm | Dosing time (DPC) | Total number of NHPs | Death times (DPC) | NHPs censored at day 28 |
|---|---|---|---|---|
| Vehicle | Not applicable | 6 | 7, 8, 8, 9, 11 | 1 |
| Treatment | DPC 1 | 6 | 9 | 5 |
| Treatment | DPC 2 | 8 | 7, 12, 13 | 5 |
| Treatment | DPC 3 | 8 | 10, 10, 11, 15 | 4 |
| Treatment | DPC 4 | 8 | 8, 9, 10 | 5 |

We fitted a pooled piecewise-exponential proportional hazards model to the reconstructed individual-level survival data. Follow-up was split into intervals defined by the unique observed death times and dosing times up to day 15, the time of the last observed death. The vehicle hazard was modelled as piecewise constant, with the baseline hazard shared across treatment arms. For each antiviral-treated arm, the treatment effect was applied only after the scheduled dosing time, so that the pre-dose hazard was identical to the vehicle hazard. The split-time likelihood was fitted using the standard Poisson representation of a piecewise-exponential survival model, with interval exposure time as an offset and a post-dose treatment multiplier applied to the baseline hazard. Baseline interval hazards were profiled out analytically, and the remaining efficacy-curve parameters were estimated by maximum likelihood.

Treatment was parameterised as an efficacy effect on the hazard scale. For an arm dosed at day $(d)$, the post-dose hazard was written as:

$$h(t \mid d) = h_0(t)\,\{1 - \mathrm{eff}(d)\}\ h(t \mid d) = h_0(t)\,\{1 - \mathrm{eff}(d)\}$$

where $(h_0(t))$ is the vehicle hazard and $(\mathrm{eff}(d))$ is the DPC-specific efficacy against death on the hazard scale. The final efficacy curve used a scaled sigmoidal function. We first defined the underlying logistic term:

$$A(x) = \frac{1}{1+\exp k(x-d_{50})}$$

where $(k)$ controls the steepness of the decline and $(d_{50})$ is the midpoint of the transition. Efficacy was then scaled to equal $(E_0)$ at DPC 0 and to reach zero at DPC 15:

$$\mathrm{eff}(d) = E_0\frac{A(d)-A(15)}{A(0)-A(15)}, \quad d < 15$$

$$\mathrm{eff}(d) = 0, \quad d \geq 15$$

$(k)$ was fixed at 1 to provide a stable sigmoidal shape given the small number of DPC arms, while $(E_0)$ and $(d_{50})$ were estimated from the pooled survival likelihood. This fitted model gave $(E_0 = 0.823)$, $(d_{50} = 5.6)$ days, and a root-mean-square error of 0.097 against empirical efficacy estimates at DPC 1–4.

Empirical DPC-specific efficacy points were calculated from the reconstructed Kaplan–Meier survival estimates using cumulative hazards:

$$H = -\log(S)$$

$$\mathrm{eff}_{emp}(d) = 1 - \frac{H_d}{H_0}$$

where $(H_d)$ is the cumulative hazard in the DPC-specific treatment arm and $(H_0)$ is the cumulative hazard in the vehicle arm. Uncertainty in these empirical points was estimated using non-parametric bootstrap resampling of animals within the vehicle and DPC-specific treatment arms. The uncertainty ribbon around the fitted sigmoid was generated from approximate model-based uncertainty in $(E_0)$ and $(d_{50})$, obtained from the Hessian of the profiled likelihood and propagated by simulation across the fitted efficacy curve **(Figure S3).**

In the transmission model, this NHP-derived hazard-scale efficacy curve was used as a delay-dependent proxy for the antiviral PEP efficacy parameter, mapping delay from exposure to first dose onto assumed prophylactic effectiveness. The curve should therefore be interpreted as a biologically informed scenario input rather than a direct estimate of human outbreak effectiveness. An important limitation is that DPC in the NHP model may not map directly onto days post-exposure in humans. In NHP challenge studies, animals are exposed to a standardised, high-dose inoculum by a defined route and typically develop viraemia and fever within a shorter and more reproducible timeframe than naturally exposed humans. A delay of several days after NHP challenge may therefore correspond to a later biological stage of infection than the same nominal delay after human exposure. For this reason, the fitted DPC curve was interpreted as a biologically informed scenario input for delayed initiation, rather than as a direct translation of NHP timing to human prophylactic effectiveness.

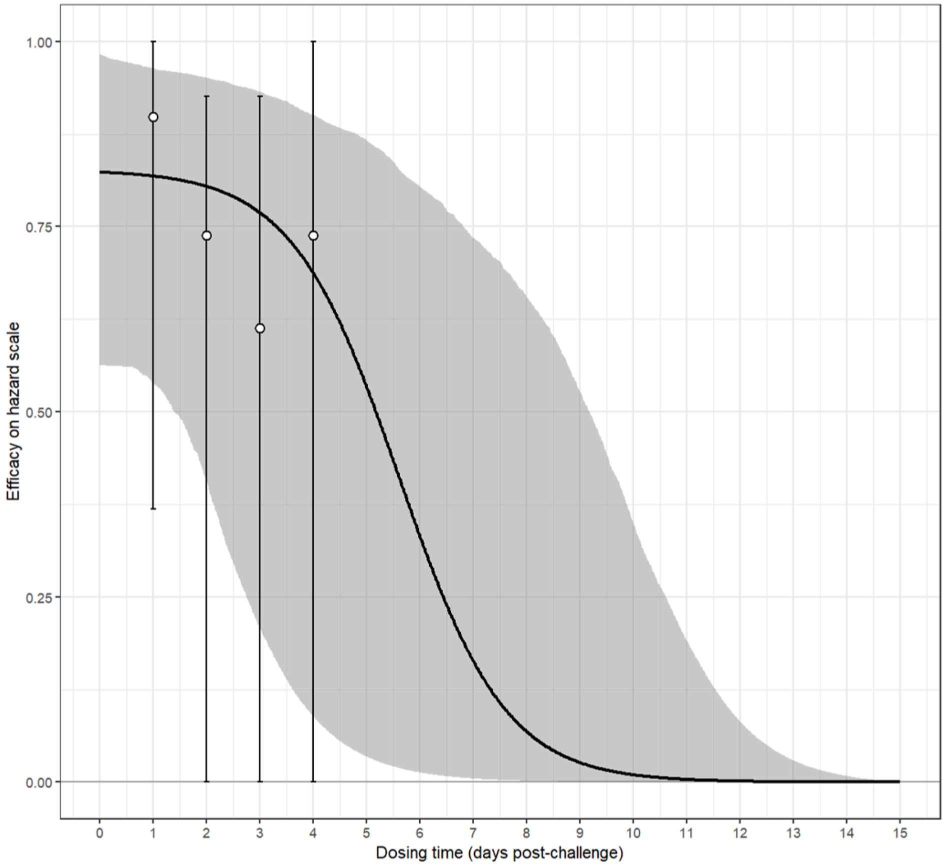


**Figure S3. NHP-informed delay-to-initiation efficacy curve.** Empirical efficacy estimates were derived from reconstructed NHP Kaplan–Meier survival data for treatment arms dosed 1–4 days post-challenge, relative to vehicle, using cumulative hazards. Points show DPC-specific empirical estimates, vertical intervals show bootstrap uncertainty, the solid line shows the fitted scaled sigmoid, and the shaded ribbon shows approximate model-based uncertainty propagated from fitted $(E_0)$ and $(d_{50})$. Efficacy was constrained to equal $(E_0)$ at DPC = 0 and zero at DPC = 15. DPC=days post-challenge; NHP=non-human primate.

To capture uncertainty in the rate at which efficacy declines with dosing delay, two additional curves were derived by applying the lower and upper bounds of the interquartile range of the shape parameter from the previously estimated logistic function, while holding median efficacy at DPC 0 constant (**Figure S4**). These bounds are carried forward in the main-text analyses as the optimistic and pessimistic efficacy scenarios, bracketing the central estimate.

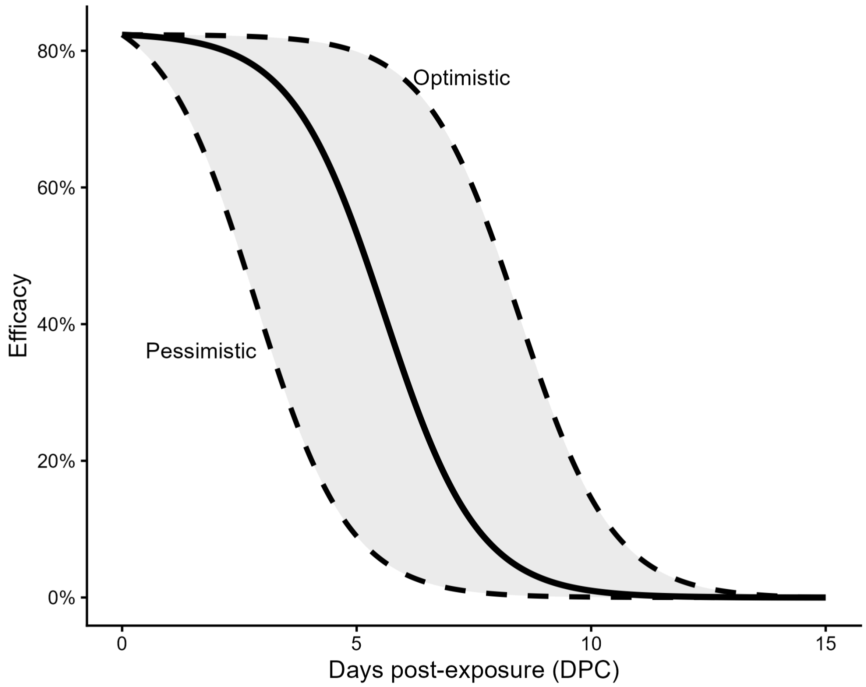


**Figure S4. Delay–efficacy relationship used to parameterise antiviral PEP in the DRC archetype operational-disruption analysis.** The solid line shows the central estimated relationship between prophylactic efficacy and the delay from HCW exposure to first antiviral dose (days post-exposure, DPC), derived from NHP survival data using a hazard-scale efficacy model. Dashed lines show the optimistic and pessimistic bounds, constructed by applying the lower and upper bounds of the interquartile range of the logistic shape parameter while holding efficacy at DPC 0 constant. The shaded region spans the optimistic and pessimistic curves. These three curves are carried forward as the central, optimistic, and pessimistic delay–efficacy scenarios in the main-text analyses and should be interpreted as biologically informed scenario inputs rather than direct estimates of human outbreak effectiveness. DPC=days post-exposure to first antiviral dose.

##### 2.5 Construction of the DRC-like operational-disruption curve

For the operational disruption analysis presented in **Figure 3**, we constructed an additional time-varying curve to represent conflict-associated deterioration in PEP delivery conditions. SDB performance data from Warsame et al.[40] were used as the primary empirical signal for operational response quality because they provide a longitudinal measure of response reach during the North Kivu and Ituri outbreak. However, SDB records capture burial alerts received by response teams and could under-represent deterioration during periods of restricted access if deaths were not reported, alerts were not generated, or teams could not reach affected communities. We therefore supplemented the SDB trajectory with community-death indicators extracted from WHO, UNICEF, DRC Ministry of Health, and response situation reports. Because these reports used heterogeneous denominators and reporting windows, community-death observations were used as qualitative anchors for the timing and relative severity of disruption rather than as a directly fitted epidemiological time series. Selected anchor points are shown in **Table S4**; the full extraction table and processing code are available in the accompanying GitHub repository.

The operational-disruption curve was constructed by hybridising the SDB performance trajectory with the community-death trend. Outside the disruption window, the SDB-derived trajectory was retained as the primary operational proxy. During the disruption window, the curve was adjusted to reflect reduced response reach indicated by increased community deaths, before recovery followed the post-disruption SDB trajectory. The resulting curve should therefore be interpreted as a scenario input representing plausible conflict-associated disruption to PEP delivery, rather than as an independently estimated measure of outbreak response performance. This hybrid curve was used only in the DRC-like operational-disruption simulations to define the timing and relative severity of reduced PEP coverage and increased delay from HCW exposure to first dose.

The hybrid operational-disruption curve was then mapped onto two PEP delivery parameters: coverage of eligible HCW exposures and DPC. Coverage was scaled linearly such that the peak of the hybrid curve corresponded to 80% coverage and the trough corresponded to 0%:

$$C(t) = 0.8 \times \frac{SDB(t)}{\max(SDB(t))}$$

DPC was scaled inversely such that the peak corresponded to a delay of 1 day and the trough to the maximum assumed delay of 5 days:

$$DPC\,(t) = 1 + 4\left(1 - \frac{SDB(t)}{\max(SDB(t))}\right)$$

To reflect that delivery conditions were not yet disrupted at outbreak onset, DPC was held at 1 day prior to the pre-conflict coverage peak. The resulting time-varying trajectories are shown in **Figure S5**.

**Table S4. Selected community-death indicators used to inform the DRC-like operational-disruption curve.** Selected community-death indicators extracted from situation reports and operational summaries for the 2018–2020 North Kivu and Ituri outbreak. Relative day is calculated from Aug 1, 2018, the date on which the outbreak was declared. Because reporting denominators and windows differed across sources, these observations were used as qualitative anchors for the timing and relative severity of operational disruption rather than as directly fitted time-series data. The complete extraction table is available in the accompanying GitHub repository.

| Period represented | Relative outbreak day | Community-death indicator | Reporting basis | Source |
|---|---|---|---|---|
| Aug 2018 | 2–14 | 61–70% | Probable cases or early reported cases used as a proxy for community deaths | 64,65 |
| Oct–Nov 2018 | 91 | 50% | Community deaths among confirmed cases in a recent reporting window | 66 |
| Dec 2018 | 127–149 | 57–64% | Community deaths among reported or confirmed cases | 67,68 |
| Feb 2019 | 191 | 71% | Peak weekly proportion of cases reported as community deaths | 69 |
| Mar–May 2019 | 236–297 | 34–55% | Community deaths among confirmed cases or recent case windows | 69,70 |
| Jun–Aug 2019 | 316–372 | 23–29% | Community deaths among confirmed cases or confirmed deaths | 71–73 |
| Nov 2019–Feb 2020 | 484–542 | 8–20% | Community deaths among confirmed cases or confirmed deaths | 74 |

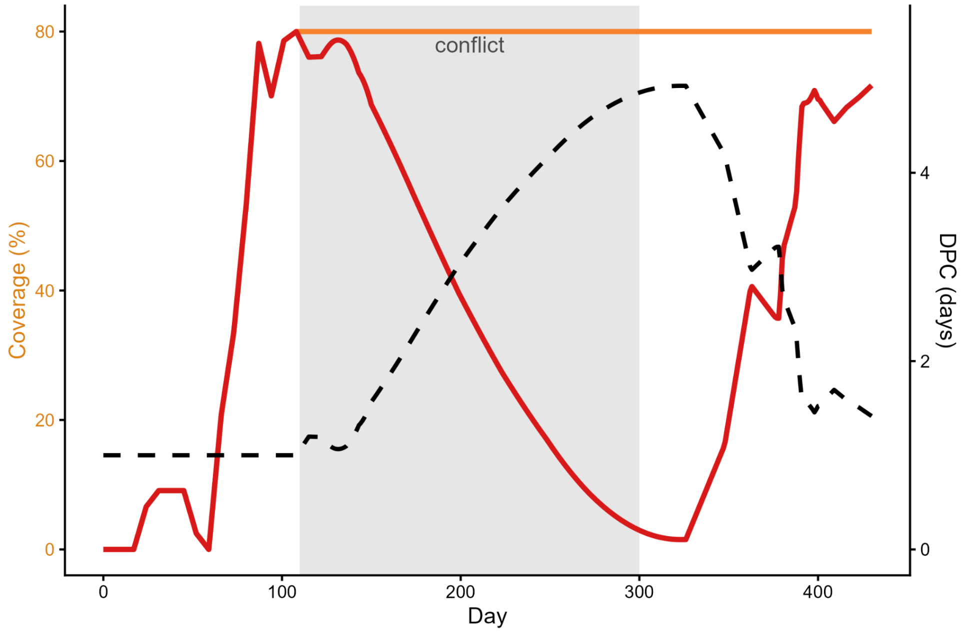


**Figure S5. Time-varying PEP coverage and dosing delay derived from the hybrid operational-disruption curve.** PEP coverage (left axis) and delay from HCW exposure to first antiviral dose (DPC, right axis) were mapped from the hybrid SDB-based operational-disruption curve (Section 2.5). The orange line shows coverage under the delayed-dosing-only scenario, in which coverage is held at its pre-conflict peak throughout. The red line shows coverage under the delayed coverage and dosing scenario, in which coverage tracks the full disruption trajectory and falls during the conflict period before partially recovering. The black dashed line shows the shared DPC trajectory under both disrupted scenarios: DPC is fixed at 1 day prior to the pre-conflict coverage peak to reflect undisrupted baseline delivery conditions. The grey shaded region indicates the simulated conflict period. DPC=days from HCW exposure to first antiviral dose; HCW=health-care worker; PEP=post-exposure prophylaxis; SDB=safe and dignified burial.

#### 3. Scenarios & Simulations Carried out

##### 3.1 Overview of simulation design

All simulation analyses used a paired direct-effect counterfactual design. For each stochastic replicate, the model was run with antiviral PEP enabled. During each run, the model recorded two linked outputs: the realised transmission tree, containing infections that occurred despite PEP, and the set of eligible HCW infections that were directly prevented by PEP. For each prevented infection, the model also reconstructed the counterfactual clinical outcome that would have occurred in the absence of PEP, using the same natural-history assumptions applied to realised infections. The with-PEP trajectory was summarised directly from the realised transmission tree. The no-PEP counterfactual was constructed by adding the infections directly prevented by PEP, and their reconstructed deaths and HCW-days lost, back to the realised outbreak. This allowed each intervention scenario to be compared with a matched no-antiviral counterfactual derived from the same stochastic replicate, reducing noise from differences in outbreak timing, size, and exposure history. This approach estimates the direct benefit of PEP for exposed HCWs. Prevented infections were not allowed to generate additional onward transmission chains in the reconstructed no-PEP counterfactual. Estimates of deaths averted and HCW-days lost averted therefore refer to direct HCW protection, rather than the total population-level effect including any secondary cases that might have followed from prevented HCW infections. This is appropriate for the primary outcomes of HCW mortality and workforce preservation, and is likely conservative for broader outbreak impact.

##### 3.2 Posterior sampling and parameter set construction

For each of the two outbreak archetypes ("West Africa-like" and "DRC-like"), 200 parameter sets (referred to throughout as posterior draws) were drawn from the ABC posterior distributions described in **Section 1.4**. These were combined with the archetype-specific fixed natural-history parameters (**Table S1**) and the posterior mean of each time-varying response trajectory (**Table S2** and **Figure S6**) to construct a complete set of model inputs for that posterior draw. The time-varying trajectories (i.e. hospitalisation probability and delay, ETU proportion, PPE/IPC coverage, and unsafe-funeral probabilities) were treated as fixed inputs conditional on the inferred response-maturation trajectory for each archetype; uncertainty in these trajectories was not propagated into the antiviral scenario analyses.

##### 3.3 Stochastic replication and takeoff conditioning

For each posterior draw and simulation setting, ten independent stochastic replicates were run, each initialised with 25 seeding infections and assigned a unique seed. This yields 2,000 stochastic replicates per setting per archetype (200 posterior draws × 10 stochastic replicates). Because a proportion of replicates will go extinct before generating a substantial outbreak, each replicate was conditionally accepted only if the realised transmission tree reached at least 100 cumulative deaths, which serves as the operational definition of outbreak takeoff throughout. If a replicate did not reach this threshold, it was re-run.

##### 3.4 Simulation settings

Simulation settings were defined by the combination of PEP coverage trajectory, intrinsic antiviral efficacy, delivery delay, stockpile size, and allocation policy, and were run separately for each outbreak archetype unless otherwise stated. We defined five sets of simulation settings corresponding to the main analyses. First, for the central immediate-availability analysis, PEP was available from day 0 with immediate coverage of eligible HCW exposures. The central illustrative assumption used 80% coverage and 80% intrinsic antiviral efficacy. Additional immediate-availability settings varied intrinsic antiviral efficacy from 50% to 90% in 10 percentage-point increments, with coverage otherwise fixed, to characterise the relationship between assumed antiviral efficacy and HCW protection. Second, for the implementation-readiness analysis, we compared three coverage trajectories. Scenario 1 ("high readiness, pre-positioned stockpiles") assumed immediate 100% coverage from day 0. Scenario 2 ("intermediate readiness, no stockpiles") used a clamped cubic spline anchored at 0% coverage on day 0, 40% on day 90, and 80% on day 180, with coverage held at 80% thereafter. Scenario 3 ("low readiness") used a clamped cubic spline that remained at 0% until day 75 and rose to 50% by day 365. Each implementation trajectory was combined with intrinsic antiviral efficacy values from 50% to 90%. Third, for the static efficacy–coverage sensitivity analysis, PEP was available from the start of the outbreak, but coverage was fixed rather than scaled up over time. We evaluated a fully factorial grid of five constant coverage levels (10%, 30%, 50%, 70%, and 90%) and five intrinsic antiviral efficacy levels (50%, 60%, 70%, 80%, and 90%), yielding 25 parameter combinations per archetype. This analysis separated the effect of limited eventual access from the effect of delayed implementation. Fourth, for the delay-to-initiation and operational-disruption analysis, we used the NHP-informed delay-to-initiation efficacy curve described in Section 2.4 and the DRC-like operational-disruption curve described in Section 2.5. These simulations were performed in the DRC-like archetype. The achievable-impact scenario assumed high PEP coverage with no delay from HCW exposure to first dose. Disruption scenarios then evaluated reduced PEP coverage, increased delay to first dose, or both, with the timing and relative severity of disruption defined by the operational-disruption curve. Central, optimistic, and pessimistic delay-to-initiation efficacy assumptions were used to assess uncertainty in the rate at which efficacy declined with dosing delay. Fifth, for the stockpile, allocation, and dose-efficiency analyses, we evaluated how finite supply, delivery delay, and allocation policy affected the number of doses required to avert HCW deaths. We compared prompt delivery, defined as same-day dosing after exposure, with delayed delivery, defined as first dose 5 days after exposure. Stockpile analyses varied the number of available doses and estimated HCW deaths averted under finite supply. Allocation analyses compared Policy A, broad allocation to all eligible HCW exposures, with Policy B, targeted allocation to high-risk residual exposures after PPE and ETU-based protection had failed. Dose-efficiency analyses varied intrinsic antiviral efficacy from 10% to 90% in 10 percentage-point increments, comparing allocation policies and delivery delays. Course requirements were converted into doses assuming 20 doses per PEP course.

##### 3.5 Outcome extraction and summary

For each stochastic simulation replicate, HCW deaths in the with-PEP setting and in the reconstructed no-antiviral counterfactual were identified among health-care workers class infections whose clinical course resulted in death. The number of HCW deaths averted was computed as the difference between counterfactual and with-PEP HCW deaths for each matched replicate, and the percentage averted was expressed relative to the counterfactual count. HCW-days lost were computed as the sum across infected HCWs of the interval from symptom onset to the end of the simulated outbreak, and the percentage of HCW-days lost averted was expressed relative to the counterfactual total for the same replicate. Posterior draw-level summaries were first obtained by averaging the ten stochastic replicates for each posterior draw; outcome distributions were then summarised as medians and 95% credible intervals across the 200 posterior draws. Weekly incident death trajectories were computed by binning event times into 7-day intervals, averaging across replicates within each posterior draw, and then computing quantiles across posterior draws.

##### 3.6 Dose requirements and allocation policies

Dose requirements for Figure 4 were calculated at the individual-replicate level. Under Policy B, representing high-risk exposure allocation, the number of prophylactic courses administered in each replicate included two groups: HCWs who received PEP but were still infected, and HCWs whose infection was prevented by PEP. This count therefore represents the number of courses required under an allocation strategy that targets exposures with substantial residual infection risk after PPE and ETU-based protection. Under Policy A, representing broad exposed-HCW allocation, we reconstructed the larger eligible exposure pool before PPE and ETU filtering by dividing the Policy B course count by the complement of the posterior-draw-specific PPE and ETU efficacy estimated in the ABC calibration. For each replicate, course requirements were converted into doses by multiplying by 20, the assumed number of doses per prophylactic course, and then divided by the number of HCW deaths averted in that replicate to obtain doses per HCW death averted. Replicates in which no HCW deaths were averted were excluded because this quantity was undefined. For each efficacy value, archetype, and policy, replicate-level dose requirements were summarised within posterior draws using medians and interquartile ranges, and then aggregated across the 200 posterior draws.

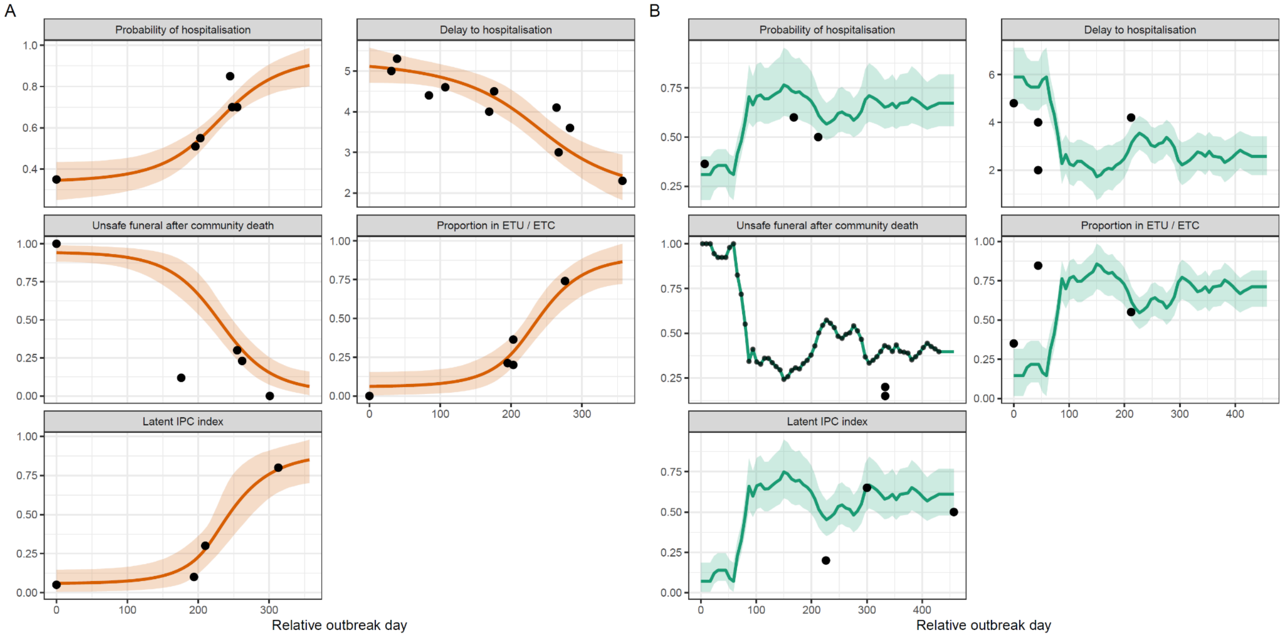


**Figure S6: Fitted time-varying response-parameter curves for the West Africa and DRC outbreak archetypes.** Time-varying response parameters used in the branching-process model were inferred from collated outbreak and literature-derived data, then converted into daily input trajectories for each archetype. **a)** Fitted response curves for the West Africa archetype, estimated using a partially pooled finite-window logistic model, and parameter-specific minimum and maximum values. **b)** Fitted response curves for the DRC archetype. For this archetype, a complete safe-and-dignified burial performance time series was used as an empirical proxy for the latent response-quality curve, and parameter-specific minimum and maximum values were estimated to map this shared curve onto each response parameter. Panels show the probability of hospitalisation, delay from symptom onset to hospitalisation, probability of an unsafe funeral after community death, proportion of hospitalized cases managed in an ETU, and a latent IPC/PPE index. Black points indicate collated observations used to inform the fit. Colored lines show posterior mean fitted trajectories, and shaded ribbons show 90% posterior credible intervals. The x-axis shows relative outbreak day, and y-axes are plotted on the natural scale of each response parameter.

### 4. Additional Results & Sensitivity Analyses

##### 4.1 Sensitivity analyses for DRC-like archetype calibration in absence of ring-vaccination

Because the DRC-like archetype was calibrated to the realised 2018–2020 North Kivu and Ituri outbreak, which included rVSV-ZEBOV-GP vaccination, we performed a sensitivity analysis to assess whether the main conclusions were robust to higher baseline transmissibility. This analysis was not intended to reconstruct ring vaccination mechanistically but provides a stress test for interpreting the DRC-like archetype in vaccine-unavailable or reduced-protection settings, where realised outbreak burden may have been larger in the absence of vaccine-derived protection. We increased baseline transmissibility in the DRC-like archetype by 10%, 20%, and 30%, while holding all other fitted parameters and time-varying response trajectories fixed. For each transmissibility level, we repeated the paired counterfactual simulations used in the primary analyses and compared baseline HCW deaths, HCW deaths averted, percentage of HCW deaths averted, and HCW-days lost averted. As expected, increasing transmissibility substantially increased baseline HCW burden. In the no-PEP counterfactual, median HCW deaths increased from 68 (95% CrI 25–155) in the fitted DRC-like archetype to 250 (173–316), 295 (238–361), and 312 (259–381) under 10%, 20%, and 30% increases in transmissibility, respectively. The absolute number of health-care workers deaths averted by PEP increased accordingly, from 54 (20–124) in the fitted archetype to 249 (206–305) under the 30% transmissibility increase. However, the proportional effect of PEP was stable across the sensitivity analysis. At 80% antiviral efficacy and full coverage, PEP averted approximately 80% of health-care workers deaths at all transmissibility levels: 80.3% (76.3–83.6) in the fitted archetype and 80.0% (78.3–81.3) under the 30% transmissibility increase. These results indicate that higher baseline transmissibility changes the absolute burden, and therefore the absolute number of deaths PEP can avert, but does not materially alter the proportional effect of PEP. They support the interpretation that the primary DRC-like results are robust, and may be conservative for higher-burden, vaccine-unavailable outbreaks.

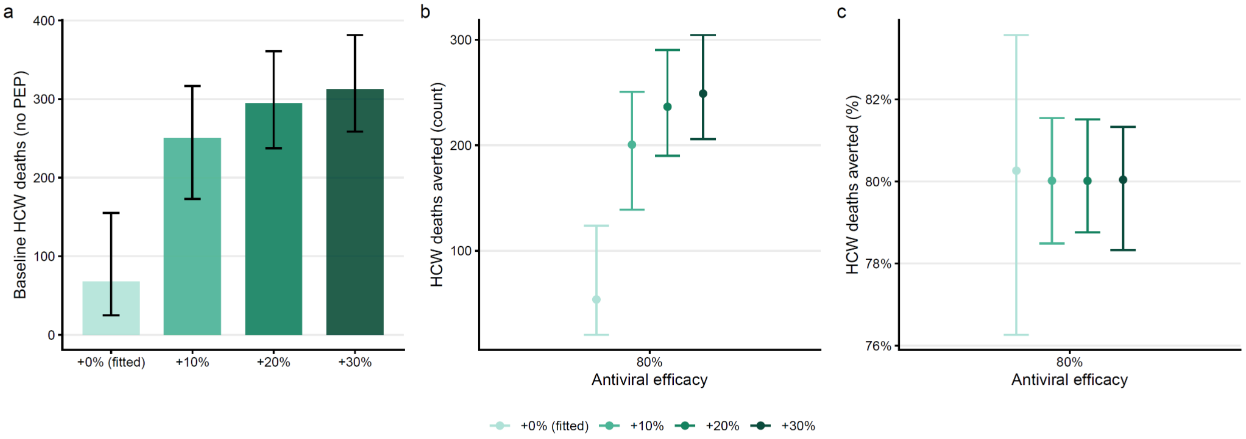


**Figure S7 Robustness of HCW-targeted PEP impact to higher transmissibility in the DRC-like archetype.** Because vaccination during the North Kivu and Ituri outbreak is likely to have suppressed transmission, the DRC-like archetype calibrated to the realised dynamics may be conservative for a setting in which a vaccine is unavailable. To test this, the DRC-like posterior simulations were rerun with baseline transmissibility increased by 10%, 20%, and 30% (by scaling the reproduction number, equivalently the per-route offspring means), holding the intervention fixed at HCW-targeted antiviral PEP with immediate 100% coverage and 80% assumed antiviral efficacy. **a)** Baseline HCW deaths in the absence of PEP, for the as-fitted archetype (+0%) and at each increased transmissibility level. **b)** Number of HCW deaths averted by PEP. **c)** Percentage of HCW deaths averted by PEP. Increasing shade indicates increasing transmissibility. Bars and points show medians; vertical error bars show 95% credible intervals across calibrated stochastic simulations. DRC=Democratic Republic of the Congo; HCW=health-care worker; PEP=post-exposure prophylaxis.

##### 4.2 Additional efficacy and coverage results

This section reports additional efficacy and coverage analyses corresponding to the immediate-availability scenarios. **Figures S7 and S9** show HCW deaths averted, while **Figures S8 and S10** show the corresponding HCW-days lost averted.

**Antiviral impact increased with assumed efficacy under immediate full coverage**
We varied assumed antiviral efficacy from 50% to 90% under an immediate full-coverage scenario, representing high PEP availability from the start of the outbreak with all eligible HCW exposures receiving PEP. Across both response archetypes, the proportion of HCW deaths averted increased approximately proportionally with assumed efficacy **(Figure S7)**. In the West Africa-like archetype, HCW deaths averted increased from 50.0% (95% CrI 48.7–55.2) at 50% efficacy to 90.0% (89.1–90.8) at 90% efficacy, with 80.0% (78.9–81.1) averted at 80% efficacy. Similar reductions were observed in the DRC-like archetype: 50.2% (45.7–55.2), 79.9% (76.1–83.8), and 90.0% (87.9–92.4) at 50%, 80%, and 90% efficacy, respectively. HCW-days lost averted followed the same pattern **(Figure S8)**. In the West Africa-like archetype, HCW-days lost averted increased from 50.0% (48.7–51.1) at 50% efficacy to 90.0% (89.2–90.7) at 90% efficacy, with 80.0% (79.0–81.1) averted at 80% efficacy. In the DRC-like archetype, the corresponding estimates were 50.0% (45.5–53.7), 80.0% (76.8–82.7), and 90.0% (87.5–92.2).

**PEP impact depended jointly on antiviral efficacy and coverage**
We next varied antiviral efficacy and coverage together to assess how incomplete coverage modified the impact of HCW-targeted PEP under immediate availability. Across both archetypes, HCW deaths averted increased with both parameters **(Figure S9)**. At 80% efficacy, increasing coverage from 10% to 90% increased HCW deaths averted from 8.0% (7.3–8.6) to 72.0% (70.5–73.6) in the West Africa-like archetype, and from 7.9% (6.1–10.3) to 72.1% (68.0–76.8) in the DRC-like archetype. At 90% coverage, increasing assumed efficacy from 50% to 90% increased HCW deaths averted from 45.1% (43.9–46.4) to 81.0% (79.8–82.1) in the West Africa-like archetype, and from 45.0% (40.5–50.6) to 81.0% (77.4–83.7) in the DRC-like archetype. Similar patterns were observed for HCW-days lost **(Figure S10)**. At 80% efficacy, increasing coverage from 10% to 90% increased HCW-days lost averted from 8.0% (7.4–8.6) to 72.0% (70.9–73.5) in the West Africa-like archetype, and from 7.9% (6.1–10.1) to 72.0% (68.5–75.8) in the DRC-like archetype. At 90% coverage, increasing assumed efficacy from 50% to 90% increased HCW-days lost averted from 45.1% (43.9–46.2) to 81.0% (80.1–82.0) in the West Africa-like archetype, and from 45.1% (40.6–48.4) to 80.8% (77.9–84.1) in the DRC-like archetype. These results show that PEP impact depends jointly on antiviral efficacy and operational coverage: high efficacy cannot compensate for poor delivery, while high coverage can preserve substantial HCW capacity even when efficacy is moderate.

##### 4.3 Sensitivity analysis for coverage ramp-up and conflict-associated disruption scenario

In the main analysis (**Figure 3**), we compared three deployment-readiness scenarios: immediate full coverage, scale-up to 80% coverage over 180 days, and delayed scale-up to 50% coverage over one year. To explore sensitivity to these assumptions, we extended this analysis across a grid of time-varying coverage trajectories defined by the onset of scale-up (0–100 days after outbreak start) and the maximum coverage attained (20–100%), with antiviral efficacy fixed at 80%. Coverage remained at zero until the specified onset day, then increased over a 180-day ramp to the specified maximum coverage (**Figure S11a**). Earlier onset and higher maximum coverage consistently increased HCW deaths averted in both archetypes (**Figure S11b,c**). Under the highest-coverage, earliest-onset trajectory, HCW deaths averted reached 75.0% (71.9–77.2%) in the West Africa-like archetype and 64.4% (49.0–72.4%) in the DRC-like archetype. Delaying onset to day 100 while maintaining 100% maximum coverage reduced these values to 47.8% (39.6–55.4%) and 48.9% (20.5–62.1%), respectively. Limiting maximum coverage to 20% with immediate onset reduced impact to 15.1% (13.9–16.0%) and 13.3% (9.5–15.6%), and combining delayed onset with 20% maximum coverage reduced impact further to 9.7% (7.5–11.5%) and 9.7% (3.8–13.6%). These results indicate that both the timing of initial availability and the achievable coverage ceiling shape PEP impact, with delays and low coverage compounding to reduce benefit. Weekly incident HCW deaths underlying the cumulative trajectories shown in **Figure 3a** are presented in **Figure S12**, stratified by efficacy assumption. Under the pessimistic assumption, a pronounced secondary surge in HCW deaths is evident during the conflict-affected period, driven by the increase in dosing delay as operational conditions deteriorate. This surge is most apparent when both coverage and dosing are disrupted, and is substantially attenuated under the optimistic efficacy assumption, reflecting the sensitivity of outbreak-period impact to the assumed rate at which prophylactic protection declines with delayed initiation.

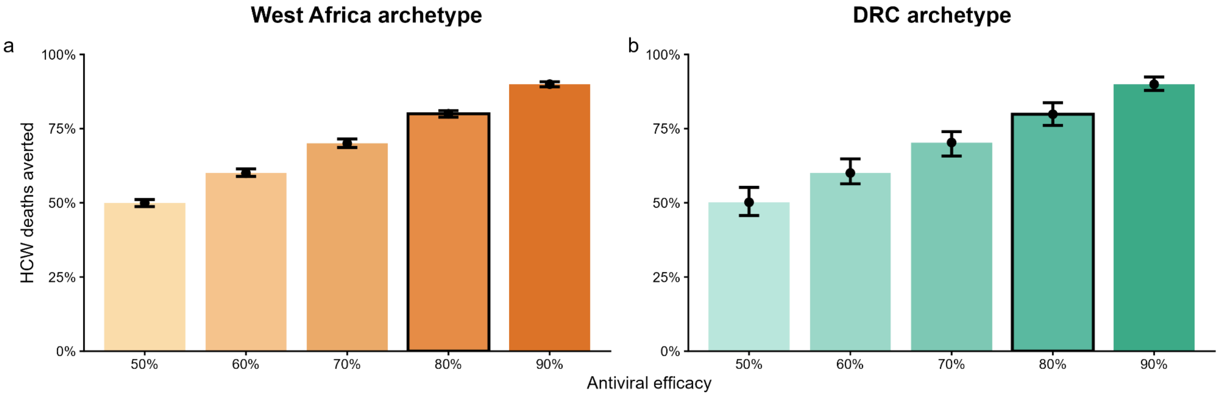


**Figure S7. Effect of antiviral efficacy on HCW deaths under immediate full-coverage PEP.** Assumed antiviral efficacy was varied from 50% to 90%, with PEP available from the start of the outbreak and delivered to all eligible HCW exposures. **a**, Percentage of HCW deaths averted in the West Africa-like archetype. **b**, Percentage of HCW deaths averted in the DRC-like archetype. The black-outlined bars indicate the 80% efficacy scenario used as the central illustrative assumption. Bars and points show medians; vertical intervals show 95% credible intervals across calibrated stochastic simulations. DRC=Democratic Republic of the Congo; HCW=health-care worker; PEP=post-exposure prophylaxis.

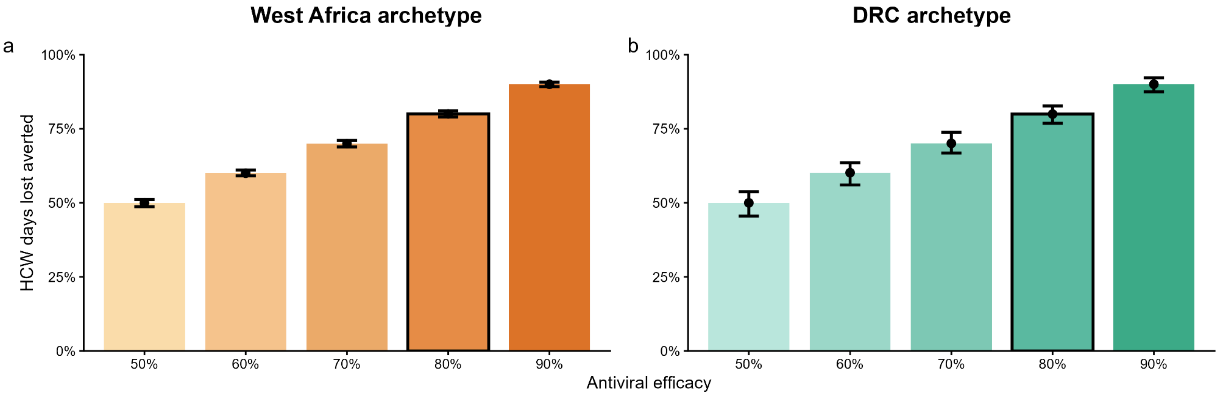


**Figure S8. Effect of antiviral efficacy on HCW-days lost under immediate full-coverage PEP.** Assumed antiviral efficacy was varied from 50% to 90%, with PEP available from the start of the outbreak and delivered to all eligible HCW exposures. a, Percentage of HCW-days lost averted in the West Africa-like archetype. b, Percentage of HCW-days lost averted in the DRC-like archetype. The black-outlined bars indicate the 80% efficacy scenario used as the central illustrative assumption. Bars and points show medians; vertical intervals show 95% credible intervals across calibrated stochastic simulations. DRC=Democratic Republic of the Congo; HCW=health-care worker; PEP=post-exposure prophylaxis.

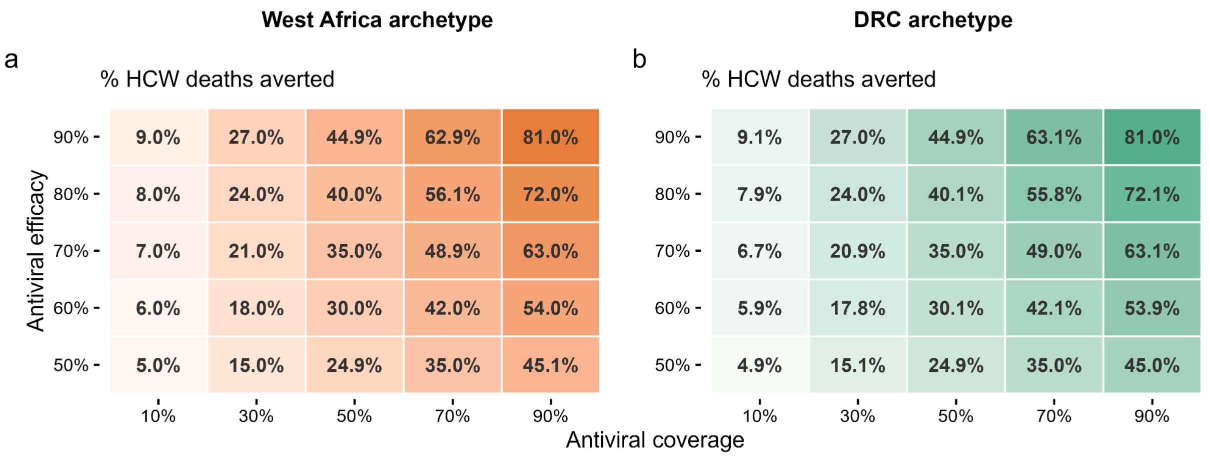


**Figure S9. Joint effect of antiviral efficacy and coverage on HCW deaths averted.** Sensitivity analysis varying assumed antiviral efficacy and coverage of eligible HCW exposures under immediate PEP availability. **a**, Percentage of HCW deaths averted in the West Africa-like archetype. **b**, Percentage of HCW deaths averted in the DRC-like archetype. Cell values show median percentages across calibrated stochastic simulations. DRC=Democratic Republic of the Congo; HCW=health-care worker; PEP=post-exposure prophylaxis.

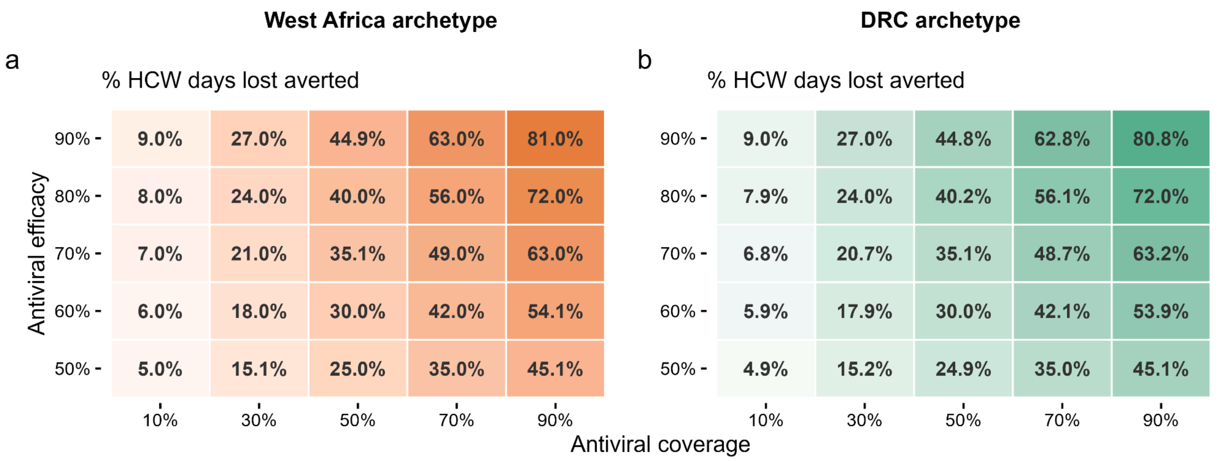


**Figure S10. Joint effect of antiviral efficacy and coverage on HCW-days lost averted.** Sensitivity analysis varying assumed antiviral efficacy and coverage of eligible HCW exposures under immediate PEP availability. **a**, Percentage of HCW-days lost averted in the West Africa-like archetype. **b**, Percentage of HCW-days lost averted in the DRC-like archetype. Cell values show median percentages across calibrated stochastic simulations. DRC=Democratic Republic of the Congo; HCW=health-care worker; PEP=post-exposure prophylaxis.

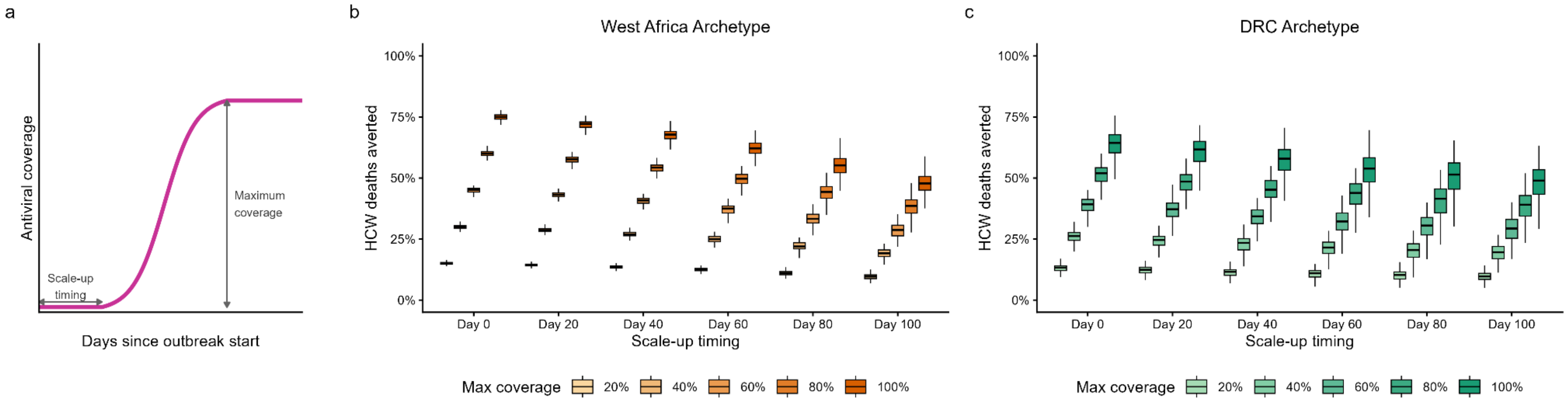


**Figure S11: Sensitivity of HCW deaths averted to scale-up timing and maximum coverage at 80% antiviral efficacy.** Antiviral PEP coverage was varied across a grid of 30 combinations defined by the onset of scale-up (0 to 100 days post-outbreak, in 20-day steps) and the maximum coverage attained (20% to 100%, in 20% steps), with antiviral efficacy fixed at 80%. Panel a illustrates how the onset of scale-up and maximum coverage jointly define the time-varying coverage trajectory used in the simulation. Panels b and c show the percentage of HCW deaths averted for the West Africa-like and DRC-like archetypes, respectively. Within each group of boxes, colours indicate maximum coverage from lightest (20%) to darkest (100%). Centre lines indicate medians, boxes indicate interquartile ranges, and whiskers indicate the full range across calibrated stochastic simulations

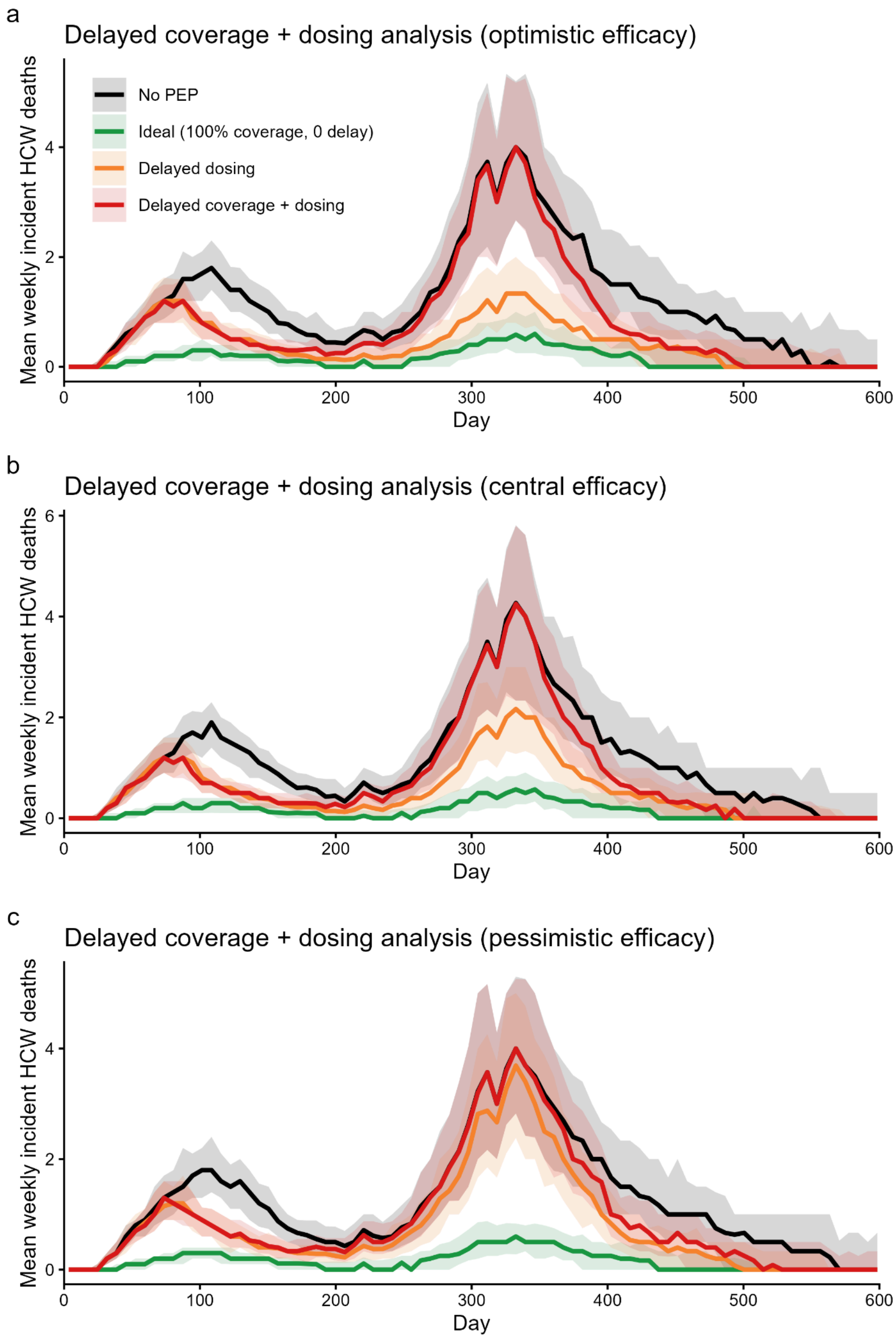


**Figure S12. Weekly incident HCW deaths under coverage and dosing disruption scenarios, by delay–efficacy assumption.** Mean weekly incident HCW deaths in the DRC-like archetype under four delivery scenarios: no PEP (black), ideal delivery with 100% coverage and no dosing delay (green), delayed dosing only with coverage preserved (orange), and combined delayed coverage plus dosing (red). Results are shown separately for optimistic (**a**), central (**b**), and pessimistic (**c**) efficacy assumptions. Shaded areas show the interquartile range across calibrated stochastic simulations. HCW=health-care worker; PEP=post-exposure prophylaxis.

**References:**


1 Sebastian Funk, Flavio Finger, James M. Azam. bpmodels: Analysing transmission chain statistics using branching process models. 2023. https://github.com/epiverse-trace/bpmodels/.

2 Hellewell J, Abbott S, Gimma A, *et al.* Feasibility of controlling COVID-19 outbreaks by isolation of cases and contacts. *Lancet Glob Health* 2020; **8**: e488–96.

3 Whittaker C, Barnsley G, Mesa DO, *et al.* Quantifying the impact of a broadly protective sarbecovirus vaccine in a future SARS-X pandemic. *Nat Commun* 2025; **16**: 8495.

4 Ebola Virus Disease in Health Care Workers — Sierra Leone, 2014. 2014; published online Dec 12. https://www.cdc.gov/mmwr/preview/mmwrhtml/mm6349a6.htm (accessed June 6, 2026).

5 Baller A, Padoveze MC, Mirindi P, *et al.* Ebola virus disease nosocomial infections in the Democratic Republic of the Congo: a descriptive study of cases during the 2018-2020 outbreak. *Int J Infect Dis* 2022; **115**: 126–33.

6 WHO Ebola Response Team, Aylward B, Barboza P, *et al.* Ebola virus disease in West Africa--the first 9 months of the epidemic and forward projections. *N Engl J Med* 2014; **371**: 1481–95.

7 Woolsey C, Cross RW, Chu VC, *et al.* The oral drug obeldesivir protects nonhuman primates against lethal Ebola virus infection. *Sci Adv* 2025; **11**: eadw0659.

8 Cross RW, Woolsey C, Prasad AN, *et al.* Oral obeldesivir provides postexposure protection against Marburg virus in nonhuman primates. *Nat Med* 2025; **31**: 1303–11.

9 International Ebola Response Team, Agua-Agum J, Ariyarajah A, *et al.* Exposure patterns driving Ebola transmission in West Africa: A retrospective observational study. *PLoS Med* 2016; **13**: e1002170.

10 Ohimain EI, Silas-Olu D. The 2013-2016 Ebola virus disease outbreak in West Africa. *Curr Opin Pharmacol* 2021; **60**: 360–5.

11 Health worker Ebola infections in Guinea, Liberia and Sierra Leone. 2015; published online May 15. https://www.who.int/publications/i/item/WHO-EVD-SDS-REPORT-2015.1 (accessed June 13, 2026).

12 Number of Ebola Cases and Deaths in Affected Countries. https://data.humdata.org/dataset/ebola-cases-2014 (accessed June 13, 2026).

13 Ebola outbreak 2018-2020- North Kivu-Ituri. https://www.who.int/emergencies/situations/Ebola-2019-drc- (accessed June 13, 2026).

14 Mukandila Kalala D, Anto F, Guure C, Ahuka S, Lubula L, Noora C. Factors predicting fatality among health-care workers infected with Ebola in the Democratic Republic of Congo, August 2018 – June 2020. *J Interv Epidemiol Public Health* 2026; **9**. DOI:10.37432/jieph-d-25-00072.

15 World Health Organization. Regional Office for Africa. Ebola Virus Disease Democratic Republic of Congo: External situation report 98. World Health Organization. Regional Office for Africa. 2020; published online June 23. https://iris.who.int/handle/10665/332654 (accessed June 13, 2026).

16 Legrand J, Grais RF, Boelle PY, Valleron AJ, Flahault A. Understanding the dynamics of Ebola epidemics. *Epidemiol Infect* 2007; **135**: 610–21.

17 Camacho A, Kucharski A, Aki-Sawyerr Y, *et al.* Temporal changes in Ebola transmission in Sierra Leone and implications for control requirements: A real-time modelling study. *PLoS Curr* 2015; **7**: ecurrents.outbreaks.406ae55e83ec0b5193e30856b9235e.

18 Faye O, Boëlle P-Y, Heleze E, *et al.* Chains of transmission and control of Ebola virus disease in Conakry, Guinea, in 2014: an observational study. *Lancet Infect Dis* 2015; **15**: 320–6.

19 Washington ML, Meltzer ML, Centers for Disease Control and Prevention (CDC). Effectiveness of Ebola treatment units and community care centers - Liberia, September 23-October 31, 2014. *MMWR Morb Mortal Wkly Rep* 2015; **64**: 67–9.

20 Polonsky JA, Böhning D, Keita M, *et al.* Novel use of capture-recapture methods to estimate completeness of contact tracing during an Ebola outbreak, Democratic Republic of the Congo, 2018-2020. *Emerg Infect Dis* 2021; **27**: 3063–72.

21 Lampaert E, Nsio Mbeta J, Nair D, *et al.* Evaluation of centralised and decentralised models of care during the 2020 Ebola Virus Disease outbreak in Equateur Province, Democratic Republic of the Congo: A brief report. *F1000Res* 2024; **13**: 642.

22 Barks PM, Camacho A, Newport T, *et al.* Evaluation of a decentralised model of care on case isolation and patient outcomes during the 2018-20 Ebola outbreak in the Democratic Republic of the Congo: a retrospective observational study. *Lancet Glob Health* 2025; **13**: e931–41.

23 Lindblade KA, Nyenswah T, Keita S, *et al.* Secondary infections with Ebola virus in rural communities, Liberia and Guinea, 2014-2015. *Emerg Infect Dis* 2016; **22**: 1653–5.

24 Tiffany A, Dalziel BD, Kagume Njenge H, *et al.* Estimating the number of secondary Ebola cases resulting from an unsafe burial and risk factors for transmission during the West Africa Ebola epidemic. *PLoS Negl Trop Dis* 2017; **11**: e0005491.

25 How to conduct safe and dignified burial of a patient who has died from suspected or confirmed Ebola or Marburg virus disease. WHO Kobe Centre. https://wkc.who.int/resources/publications/i/item/WHO-EVD-Guidance-Burials-14.2 (accessed June 14, 2026).

26 Althaus CL. Ebola superspreading. *Lancet Infect Dis* 2015; **15**: 507–8.

27 Toth DJA, Gundlapalli AV, Khader K, *et al.* Estimates of outbreak risk from new introductions of Ebola with immediate and delayed transmission control. *Emerg Infect Dis* 2015; **21**: 1402–8.

28 Ajelli M, Parlamento S, Bome D, *et al.* The 2014 Ebola virus disease outbreak in Pujehun,

Sierra Leone: epidemiology and impact of interventions. *BMC Med* 2015; **13**: 281.

29 Lau MSY, Gibson GJ, Adrakey H, *et al.* A mechanistic spatio-temporal framework for modelling individual-to-individual transmission-With an application to the 2014-2015 West Africa Ebola outbreak. *PLoS Comput Biol* 2017; **13**: e1005798.

30 Nash RK, Bhatia S, Morgenstern C, *et al.* Ebola virus disease mathematical models and epidemiological parameters: a systematic review. *Lancet Infect Dis* 2024; **24**: e762–73.

31 Velásquez GE, Aibana O, Ling EJ, Diakite I, Mooring EQ, Murray MB. Time from infection to disease and infectiousness for Ebola virus disease, a systematic review. *Clin Infect Dis* 2015; **61**: 1135–40.

32 Lindblade KA, Kateh F, Nagbe TK, *et al.* Decreased Ebola transmission after rapid response to outbreaks in remote areas, Liberia, 2014. *Emerg Infect Dis* 2015; **21**: 1800–7.

33 WHO Ebola Response Team, Agua-Agum J, Ariyarajah A, *et al.* West African Ebola epidemic after one year--slowing but not yet under control. *N Engl J Med* 2015; **372**: 584–7.

34 Guetiya Wadoum RE, Sevalie S, Minutolo A, *et al.* The 2018-2020 Ebola outbreak in the Democratic Republic of Congo: A better response had been achieved through inter-state coordination in Africa. *Risk Manag Healthc Policy* 2021; **14**: 4923–30.

35 UN Mission for Ebola Emergency Response. Global Ebola Response - Making a Difference: Progress Report 2015. UN Mission for Ebola Emergency Response, 2015 https://reliefweb.int/report/sierra-leone/global-ebola-response-making-difference-progress-report-2015-0 (accessed June 13, 2026).

36 Kilmarx PH, Clarke KR, Dietz PM, *et al.* Ebola virus disease in health care workers--Sierra Leone, 2014. *MMWR Morb Mortal Wkly Rep* 2014; **63**: 1168–71.

37 Grinnell M, Dixon MG, Patton M, *et al.* Ebola virus disease in health care workers--Guinea, 2014. *MMWR Morb Mortal Wkly Rep* 2015; **64**: 1083–7.

38 Selvaraj SA, Lee KE, Harrell M, Ivanov I, Allegranzi B. Infection rates and risk factors for infection among health workers during Ebola and Marburg virus outbreaks: A systematic review. *J Infect Dis* 2018; **218**: S679–89.

39 Curran KG, Gibson JJ, Marke D, *et al.* Cluster of Ebola Virus Disease linked to a single funeral - Moyamba district, Sierra Leone, 2014. *MMWR Morb Mortal Wkly Rep* 2016; **65**: 202–5.

40 Warsame A, Eamer G, Kai A, *et al.* Performance of a safe and dignified burial intervention during an Ebola epidemic in the eastern Democratic Republic of the Congo, 2018-2019. *BMC Med* 2023; **21**: 484.

41 Gleason B, Redd J, Kilmarx P, *et al.* Establishment of an Ebola treatment unit and laboratory - Bombali District, Sierra Leone, July 2014-January 2015. *MMWR Morb Mortal Wkly Rep* 2015; **64**: 1108–11.

42 Barbarossa MV, Dénes A, Kiss G, Nakata Y, Röst G, Vizi Z. Transmission dynamics and final epidemic size of Ebola Virus Disease outbreaks with varying interventions. *PLoS One* 2015; **10**: e0131398.

43 Ministry of Health National Coordination Commitee. National plan for the response to the ebola virus disease epidemic in north kivu province. Ministry of Health, 2018 https://www.who.int/docs/default-source/documents/spr-ebola-2019/srp1-drc-ebola-disease-outbreak-response-plan.pdf?sfvrsn=40799796_4.

44 https://www.who.int/docs/default-source/documents/emergencies/drc-ebola-response-srp-1-3-october2019.pdf (accessed June 13, 2026).

45 Rivers CM, Lofgren ET, Marathe M, Eubank S, Lewis BL. Modeling the impact of interventions on an epidemic of Ebola in Sierra Leone and Liberia. arXiv [q-bio.PE]. 2014; published online Sept 16. DOI:10.48550/arXiv.1409.4607.

46 Drake JM, Bakach I, Just MR, O'Regan SM, Gambhir M, Fung IC-H. Transmission models of historical Ebola outbreaks. *Emerg Infect Dis* 2015; **21**: 1447–50.

47 World Health Organization. Ebola response roadmap - Situation report update 26 November 2014. World Health Organization, 2014 https://reliefweb.int/report/liberia/ebola-response-roadmap-situation-report-update-26-november-2014 (accessed June 13, 2026).

48 https://reliefweb.int/report/sierra-leone/ebola-response-roadmap-situation-report-update-3-december-2014 (accessed June 13, 2026).

49 Ebola Epidemic — Liberia, March–October 2014. 2014; published online Nov 14. https://www.cdc.gov/mmwr/preview/mmwrhtml/mm63e1114a4.htm (accessed June 13, 2026).

50 https://iris.who.int/bitstream/handle/10665/136020/roadmapsitrep_8Oct2014_eng.pdf (accessed June 13, 2026).

51 World Health Organization. Ebola response roadmap - Situation report update 24 December 2014. World Health Organization, 2014 https://reliefweb.int/report/sierra-leone/ebola-response-roadmap-situation-report-update-24-december-2014 (accessed June 13, 2026).

52 Cooper C, Fisher D, Gupta N, MaCauley R, Pessoa-Silva CL. Infection prevention and control of the Ebola outbreak in Liberia, 2014-2015: key challenges and successes. *BMC Med* 2016; **14**: 2.

53 Pathmanathan I, O'Connor KA, Adams ML, *et al.* Rapid assessment of Ebola infection prevention and control needs —. https://www.cdc.gov/mmwr/pdf/wk/mm63e1209a3.pdf (accessed June 13, 2026).

54 Lewnard JA, Ndeffo Mbah ML, Alfaro-Murillo JA, *et al.* Dynamics and control of Ebola virus transmission in Montserrado, Liberia: a mathematical modelling analysis. *Lancet Infect Dis* 2014; **14**: 1189–95.

55 Nyenswah TG, Fallah M, Calvert GM, *et al.* Cluster of Ebola virus disease, bong and montserrado counties, Liberia. *Emerg Infect Dis* 2015; **21**: 1253–6.

56 Biedron C, Lyman M, Stuckey MJ, *et al.* Evaluation of infection prevention and control readiness at frontline health care facilities in high-risk districts bordering Ebola virus disease-affected areas in the Democratic Republic of the Congo - Uganda, 2018. *MMWR Morb Mortal Wkly Rep* 2019; **68**: 851–4.

57 Kabego L, Kourouma M, Ousman K, *et al.* Impact of multimodal strategies including a pay for performance strategy in the improvement of infection prevention and control practices in healthcare facilities during an Ebola virus disease outbreak. *BMC Infect Dis* 2023; **23**: 12.

58 Nielsen CF, Kidd S, Sillah ARM, *et al.* Improving burial practices and cemetery management during an Ebola virus disease epidemic - Sierra Leone, 2014. *MMWR Morb Mortal Wkly Rep* 2015; **64**: 20–7.

59 DSpace. https://iris.who.int/items/82f9c672-5800-46bc-a3a1-426001e1a355 (accessed June 13, 2026).

60 https://iris.who.int/bitstream/handle/10665/145198/roadmapsitrep_10Dec2014_eng.pdf?sequence=1 (accessed June 13, 2026).

61 https://iris.who.int/bitstream/handle/10665/149314/roadmapsitrep_21Jan2015_eng.pdf (accessed June 13, 2026).

62 Checchi F, Eamer G, Katshitshi J, Robles Dios L, Kai A, Warsame A. Effect of a safe and dignified burial intervention on Ebola virus transmission in the eastern Democratic Republic of the Congo, 2018-19: a propensity score analysis. *Lancet Glob Health* 2025; **13**: e1617–26.

63 Stan Reference Manual. Stan Docs. https://www.google.com/url?q=https://mc-stan.org/docs/reference-manual/&sa=D&source=docs&ust=1781398161590544&usg=AOvVaw2I1UfNt_9HEbkTv1a_LpHd (accessed June 13, 2026).

64 Ebola virus disease – Democratic Republic of the Congo. https://www.who.int/emergencies/disease-outbreak-news/item/4-august-2018-ebola-drc-en (accessed June 19, 2026).

65 Ebola virus disease – Democratic Republic of the Congo. https://www.who.int/emergencies/disease-outbreak-news/item/17-august-2018-ebola-drc-en (accessed June 19, 2026).

66 https://www.afro.who.int/sites/default/files/2018-11/WCO (accessed June 19, 2026).

67 EBOLA RDC - Evolution de la riposte contre l’épidémie d'Ebola dans les provinces du Nord Kivu et de l'Ituri au Mercredi 19 décembre 2018. http://translate.google.com/translate?hl=auto&langpair=auto%7Cen&u=https://us13.campaign-archive.com/%3Fu%3D89e5755d2cca4840b1af93176%26id%3Dc2608bba60 (accessed June 19, 2026).

68 https://iris.who.int/bitstream/handle/10665/277472/SITREP-EVD-DRC-20190103-eng.pdf?isAllowed=y (accessed June 19, 2026).

69 Ebola virus disease – Democratic Republic of the Congo. https://www.who.int/emergencies/disease-outbreak-news/item/16-may-2019-ebola-drc-en (accessed June 19, 2026).

70 Ebola virus disease – Democratic Republic of the Congo. https://www.who.int/emergencies/disease-outbreak-news/item/06-june-2019-ebola-drc-en (accessed June 19, 2026).

71 https://www.unicef.org/media/81086/file/DRC-Ebola-situation-reports-January-June-2019.pdf (accessed June 19, 2026).

72 https://www.unicef.org/media/81091/file/DRC-Ebola-situation-reports-July-August-2019.pdf (accessed June 19, 2026).

73 DRC Ebola situation reports from 2019. https://www.unicef.org/documents/drc-ebola-situation-reports-2019 (accessed June 19, 2026).

74 Ebola virus disease – Democratic Republic of the Congo. https://www.who.int/emergencies/disease-outbreak-news/item/2020-DON243 (accessed June 19, 2026).
